# Risk factors associated with respiratory infectious disease-related presenteeism: a rapid review

**DOI:** 10.1101/2021.04.12.21255302

**Authors:** Sarah Daniels, Hua Wei, Yang Han, Heather Catt, David W. Denning, Ian Hall, Martyn Regan, Arpana Verma, Carl A. Whitfield, Martie van Tongeren

**Author notes:** **Correspondence:** Sarah Daniels, The University of Manchester, Ellen Wilkinson Building, Oxford Road, Manchester, M13 9PL.

## Abstract

**Background:** Workplace transmission is a significant contributor to severe acute respiratory syndrome coronavirus 2 (SARS-CoV-2) outbreaks. Previous studies have found that infectious illness presenteeism could contribute to outbreaks in occupational settings and identified multiple occupational and organisational risk factors. Amid the COVID-19 pandemic, it is imperative to investigate presenteeism particularly in relation to respiratory infectious disease (RID). Hence, this rapid review aims to determine the prevalence of RID-related presenteeism, including COVID-19, and examines the reported reasons and associated risk factors.

**Methods:** The review followed a Preferred Reporting Items for Systematic Reviews and Meta-Analyses (PRISMA) search approach and focused on studies published in English and Chinese. Database searches included MEDLINE, EMBASE, Web of Science, China Knowledge Resource Integrated Database (CNKI) and preprint databases MedRxiv and BioRxiv.

**Results:** The search yielded 54 studies, of which four investigated COVID-19-related presenteeism. Prevalence of work presenteeism ranged from 14.1% to 55% for confirmed RID, and 6.6% to 100% for those working with suspected or subclinical RID. The included studies demonstrated that RID-related presenteeism is associated with occupation, sick pay policy, age, gender, health behaviour and perception, vaccination, peer pressure and organisational factors such as presenteeism culture.

**Conclusions:** This review demonstrates that presenteeism or non-adherence to isolation guidance is a real concern and can contribute to workplace transmissions and outbreaks. Policies which would support workers financially and improve productivity, should include a range of effective non-pharmaceutical inventions such as workplace testing, promoting occupational health services, reviewing pay and bonus schemes and clear messaging to encourage workers to stay at home when ill. Future research should focus on the more vulnerable and precarious occupational groups, and their inter-relationships, to develop comprehensive intervention programs to reduce RID-related presenteeism.

## BACKGROUND

The COVID-19 pandemic is changing the landscape of global public health, social and work practice in an unprecedented manner, with many workplaces employing essential infection prevention control (IPC) measures to reduce the spread of severe acute respiratory syndrome coronavirus 2 (SARS-CoV-2).

The three transmission mechanisms of SARS-CoV-2, namely contact and droplet, airborne and fomite transmission, present significant challenges to workplace disease control [1]. As a result, the overall effectiveness of workplace COVID-19 control measures often relies on workforce management policies, including isolation and “ stay at home” behaviour. The propensity for workers to enter the workplace with COVID-19 may undermine their effectiveness.

Previous reviews on transmission of infectious diseases within workplaces, including diseases impacting the gastrointestinal tract, have found that multiple occupational and organisational characteristics could contribute to infectious illness presenteeism [2]. However, reviews of existing evidence focusing solely on presenteeism and workplace transmission of respiratory infectious diseases (RIDs), including COVID-19, are lacking. Although there have been reviews addressing behavioural drivers of presenteeism in general [3], it is necessary to conduct a review focusing on RID-related presenteeism because many RIDs do not incapacitate patients immediately and thus are often perceived as minor or common diseases. This is particularly pertinent in the context of COVID-19, where the majority of working age individuals experience only mild symptoms, and yet presenteeism can have severe public health consequences.

Current studies have identified that infection rates of COVID-19 and other RIDs are higher among occupations that involve frequent social interaction and proximity with clients and co-workers [4, 5]. Previous research reported that workers may be disproportionately vulnerable to compliance failure with control measures during an influenza pandemic because of job insecurity and financial problems associated with missing work [6, 7]. More recently, low rates of self-isolation behaviours were reported in key worker sectors during the COVID-19 pandemic, likely due to greater financial need, social pressure to attend work or inability to work from home [8].

Understanding and mitigating against the motivations as to why people attend work with COVID-19, or other RID, is key in implementing effective infection control measures. This review aims to highlight the evidence for reasons and risk factors associated with presenteeism in workers with RID, including COVID-19. With the purpose of identifying potential workplace policies to encourage workers to stay at home when ill. Thus, our research questions are as follows:

*Main research question: What reasons and risk factors are associated with presenteeism in workers with RID?*

*Sub-question: what is the prevalence of RID-related presenteeism?*

## METHODS

A review protocol was pre-published on PROSPERO (ID: CRD42020224518). The review is reported in line with the Preferred Reporting Items for Systematic Reviews and Meta-Analyses (PRISMA) guidelines and adjustments made to accommodate qualitative research [9].

### Definitions

‘Presenteeism’ is commonly defined as people who attend work while ill [10]. Though frequently measured as prevalence from an epidemiological perspective, i.e. the percentage of workers who attended work while ill, it can be measured as productivity loss from a health economics perspective, i.e. the number of hours or days worked with compromised productivity due to the illness, with some converted into economic loss. In this review, focus is on prevalence and five types of presenteeism behaviour:

1. Working with an RID infection (confirmed by clinical diagnosis or laboratory testing).

2. Working with RID symptoms (suspected or subclinical).

3. Going to work with a history of exposure to RID.

4. Non-adherence to guidance to stay at home from work with infected, suspected or exposed to RID.

5. Propensity (i.e. the inclination or tendency), to attend the workplace with confirmed, suspected or exposure to RID, evaluated by hypothetical questions such as “ would you attend work whilst ill?”.

Houghton et al [11] defined RIDs as diseases that cause acute respiratory tract infection (RTI) and severe respiratory disease in susceptible people with apparently normal immune systems.

### Study selection criteria

Population: working women and men, any age.

Exposure: confirmed or suspected RID or close contact with confirmed or suspected cases (i.e. family members or shared accommodations).

Comparator: none.

Outcomes

1. Prevalence of presenteeism in the following sub-populations:
  a. workers attending work with confirmed, suspected or exposure to RID.
  b. propensity to attend the workplace with confirmed, suspected, or exposure to RID.
  c. adherence to guidance (e.g. government or physician) to stay at home from work with confirmed, suspected or exposure to RID.
2. Reported reasons for presenteeism in any of the three sub-populations listed in 1. (a-c).
3. Statistical risk factors associated with attending work in any of the three sub-populations listed in 1. (a-c).

Study design: We searched for randomised controlled trials, cohort studies, case-control studies, cross-sectional studies and case reports. Reviews, editorials, protocols and conference papers were excluded.

Language: English or Chinese.

Publication period: no restrictions.

### Review process

Two authors (SD and HW) tested the screening process with 20% of search results for all English databases to ensure consistency in the screening process. Two authors screened all the English (SD and HW) and Chinese (HW and YH) articles at title and abstract screening and full paper review stages. Differences were discussed and reconciled with input from additional authors (HC and MvT) if required.

### Information sources

We searched MEDLINE, EMBASE, PsycInfo, the Cochrane Library, Web of Science and the World Health Organization COVID-19 database for English publications, and the China Knowledge Resource Integrated Database (CNKI) for Chinese publications. We also searched the preprint databases MedRxiv and BioRxiv. For grey literature, we conducted searches on the following databases: the Public Health England COVID-19 rapid reviews database, the European Centre for Disease Prevention and Control database, the Centres for Disease Prevention and Control database and the Chinese Centre for Disease Control and Prevention. We conducted hand searches of the reference list of included studies and some excluded studies including systematic reviews. All searches were completed in March 2021.

### Search strategy

English language searches were conducted by two researchers (SD and HW). The search strategy was developed based on published reviews using similar terms, with modifications that were deemed appropriate for the purpose of this review. Specifically, we drew search terms for respiratory infectious diseases from Houghton et al [11] and infectious illness presenteeism Webster et al [2]. Search terms used for presenteeism were ‘presenteeism’, ‘going to work while ill/sick’, ‘suspected, subclinical or mild symptoms’, ‘non-compliance or violating guideline/guidance/protocol’ and ‘exposed to or contact with confirmed/diagnosed case’. We also included terms such as ‘isolation’, ‘quarantine’ ‘social distance’ or ‘lockdown’. For RID and COVID-19 diseases, we used ‘COVID’, ‘coronavirus’, ‘nCoV’, ‘SARS’, ‘MERS’, ‘flu/influenza/influenza-like’, ‘respiratory infectious disease’ and ‘respiratory tract infection’. Search terms were translated into Chinese by two Chinese speaking researchers (HW and YH) and search strategies were adapted for CNKI.

Different search strategies were trialled with consideration for both specificity and sensitivity. HW and SD carried out preliminary searches on different databases testing a variety of search strategies. These were finalised in discussions with HC and MvT. Our final search strategy used terms and associated words for ‘COVID-19’ or ‘respiratory infectious diseases’ and ‘presenteeism’, joined by the AND function. A copy of our search strategy in MEDLINE is included as Additional file 1.

### Data extraction

Data from the final set of studies was extracted by SD and HW using a data extraction table agreed by all reviewers. Data extracted included citation, study design, objectives, sample size, population, and results regarding the prevalence of RID-related presenteeism and reported reasons or statistical risk factors associated with it.

### Quality assessment

Two authors (SD and HW) assessed the risk of bias for each study independently. Any disagreements were resolved by discussion or by involving another author (MvT). The Newcastle-Ottawa Scale (NOS) was used for the longitudinal cohort studies and utilises a ‘star system’ in which each study is judged on three broad perspectives: the selection of the study groups; the comparability of the groups; and the ascertainment of outcome of interest. A modified NOS developed for previous research [12] was used for the cross-sectional studies, with questions adjusted for the assessment of studies that measure outcomes at one point in time rather than chronologically. The Critical Appraisal Skills Programme (CASP) checklists for qualitative studies uses ten items grouped into three broad issues: the validity of the study results; the data analysis process and ethical considerations; the contribution the study makes to existing knowledge or understanding. For both NOS and CASP, the subscale items were used as a tool to help evaluate the internal validity for each included study and to categorise the study quality as low, moderate or high.

### Data synthesis and analysis

Study designs and outcome measures of the included literature were heterogeneous. Consequently, we used narrative synthesis for data analysis rather than meta-analyses. The studies were grouped into the four types of RID-related presenteeism behaviour. The reported reasons and risk factors for RID-related presenteeism were structured into over-arching themes related to work factors (occupation type, work and employment, social norms and expectations, and organisational factors) and individual factors (sociodemographics, health behaviours or perception and vaccination uptake). Data synthesis and analysis was performed by SD and HW.

## RESULTS

### Search results

Our initial search yielded 794 papers after deduplication, with an additional 13 papers identified through reference list searches. After title and abstract screening, 65 papers were taken to full text review. Of these, 54 papers were selected for inclusion for data extraction. Three of the included studies were published in Chinese and 51 studies were English. See Figure 1 for a PRISMA flow diagram of the process and reasons for exclusion.

**Figure 1:**
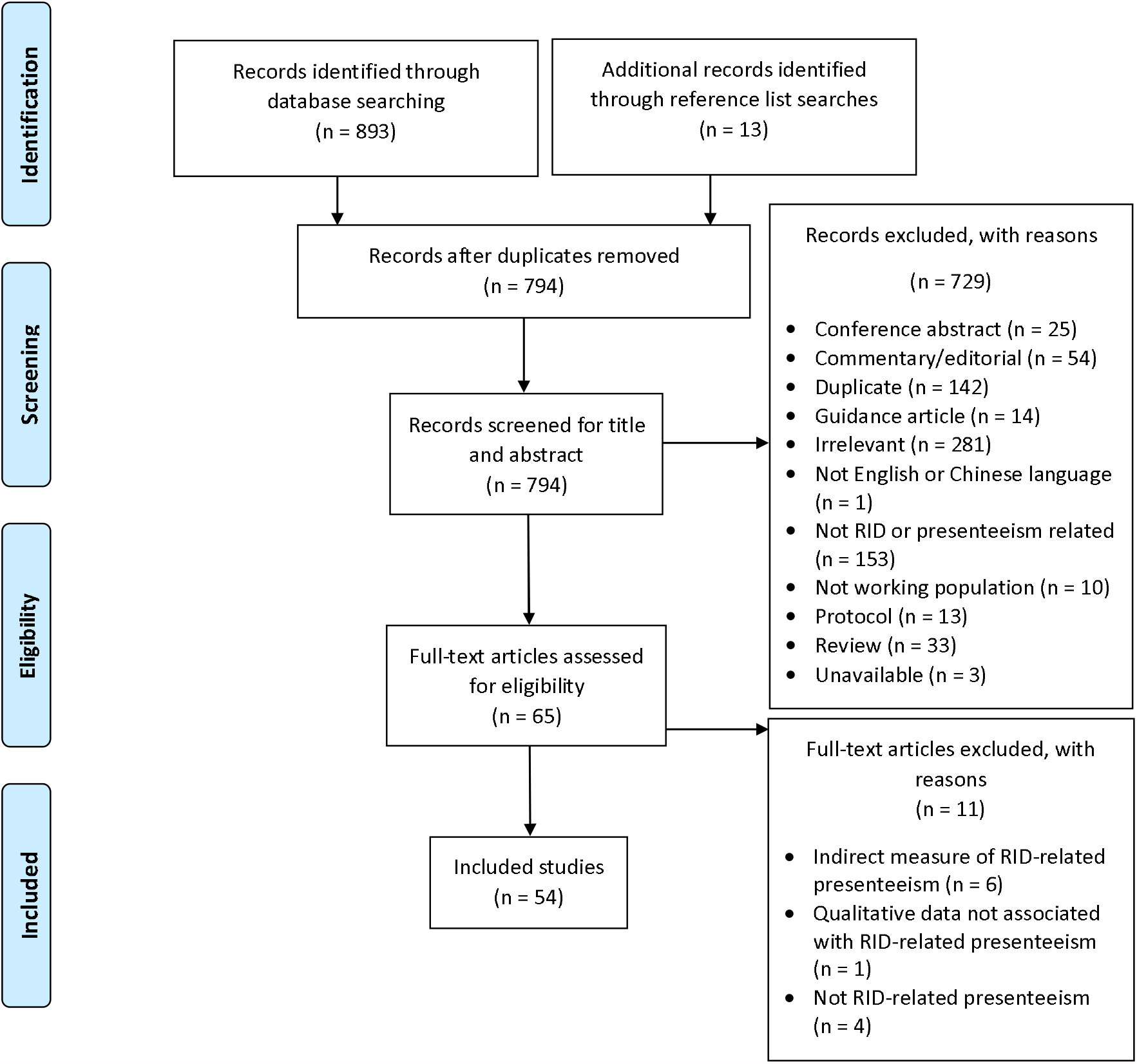
PRISMA diagram of the screening process

### Study characteristics

Of the 54 included studies, 44 were cross-sectional studies (see Additional file 2). Study locations were North America (n = 32) [6, 13–43], Asia (n = 9) [44–52], Europe (n = 7) [53–59], Australia (n = 4) [60–63], worldwide population (n = 1) [64], and an unspecified location (LinkedIn members) (n = 1) [65].

The sample size ranged from 14 to 550,360, ages ranged from 18 to >65 years old, and the percentage of females ranged from 22.5% to 99.2%. Included studies focused on healthcare occupations (n = 30) [15–20, 24, 25, 28, 29, 32, 33, 35–42, 44–46, 49, 51, 56, 59–61, 64], other specified occupations (n = 10) [14, 22, 23, 26, 27, 30, 34, 47,48,57] or general working populations (n = 14) [6, 13, 21, 31,43, 50, 52–55, 58, 62, 63, 65]. Studies investigated influenza-and/or ILI-related presenteeism (n = 38) [6, 16–28,30,31, 33, 35,36,39,41, 42, 44–48,50, 51, 54, 57, 59, 60, 62–65], general upper respiratory infection (n = 10) [13, 29, 32, 34, 37, 38, 40, 49, 55, 61], common cold or RID-type symptoms such as cough and sore throat (n = 7) [23, 36, 47, 57, 58, 62, 65], COVID-19 (n = 4) [14, 43, 52, 53], breath infections (n = 1) [56] and streptococcal infection (n = 1) [15].

### Quality assessment

The overall quality of the cross-sectional studies ranged from poor to moderate (see Additional file 3). Six studies did not describe the assessment of exposure to RID or used a survey with hypothetical questions such as “ would you attend work whilst ill?” [6, 26, 37, 48, 62, 63]. Fourteen were not representative of the target population due to recruitment from selected groups or organisations [20, 26, 28, 29, 32, 33, 38, 39, 41, 47, 51, 57, 64, 65] and only six had justified sample sizes [6, 21, 25, 45, 62, 63]. In addition, only nine studies compared respondents with non-respondents and weighted the data to the population distribution to avoid response bias [6, 17, 19, 28, 45, 57, 58, 62, 63].

The quality of the seven cohort studies ranged from moderate to high. Six studies were truly or somewhat representative of the target population [27, 34, 40, 53, 59, 60] with one study having invited participation from a selected group [30]. Only three cohort studies used formal records to ascertain the exposure to an RID. Imai et al [60] and Kuster et al [59] acquired laboratory-confirmed influenza data, and Jain et al [53] obtained data from the London Coronavirus Response Centre. No studies demonstrated that RID-presenteeism was not present at the start of the study. Four studies followed participants over an influenza season to account for the disease of interest [27, 30, 34, 59]. Of the five prospective cohort studies, only one study had a low follow-up rate (23.8%) [40].

Of the three qualitative studies, two were assessed to be moderate [14, 35] and one was of high quality [61]. The methodology and research design of the two studies, assessed to be of moderate quality, was unclear and neither study included a statement related to ethical approval. None of the qualitative studies indicated whether the researcher critically examined their own role, leaving potential bias and influence during analysis and selection of data for presentation. Only Mitchell and Coatsworth [61] gave an in-depth description of the data analysis process.

### Presenteeism measures

Varying measurement methods and recall periods were used to quantify presenteeism. Forty-six studies reported frequency of RID-related presenteeism which included prevalence and mean days/hours worked while ill, and five studies measured presenteeism as workplace productivity levels (see Additional file 4).

Six studies reported that prevalence of presenteeism ranged from 14.1% to 55% for respondents with RID confirmed by laboratory test or clinical diagnosis [15, 16, 53–55,60]. Prevalence ranged from 6.6% to 100% for symptoms of RID (suspected or subclinical) in 27 studies [17–20, 22–29, 31, 32, 39–46, 49, 51, 52, 59, 61]. For history of exposure, the prevalence was 77% for trainee physicians [33]. The propensity to attend work while ill ranged from 14% to 100% [36–38, 48, 50, 58, 64]. While 50.7% to 96.6% reported that they would adhere to guidance to stay at home from work with RID [6, 62, 63]. Within this same sub-group, a high proportion (94-96.1%) hypothesised they would stay at home with RID for at least 7 days during a confirmed pandemic.

### Reasons and risk factors for RID-related presenteeism

We grouped presenteeism reasons and risk factors into themes by work and individual factors (see Additional files 5 and 6). Reasons are defined in this context as qualitative findings concluded from answers to questions such as “ what are the main reasons that you worked while ill in the last week?” during surveys or interviews. Conversely, risk factors are based on statistically analysed associations or correlations. We recognise that reasons for presenteeism often interlink and overlap. In these cases, we have tried to assign them to the category with the best fit.

#### Occupation type

Five studies reported a sense of duty or professional obligation as reasons for RID-related presenteeism, particularly in healthcare workers and school employers [18, 19, 22, 39, 61]. The percentage of participants who chose it as one of the main (e.g. one of the top 4) reasons for presenteeism ranged from 28% to 56% within these five studies.

Eight studies measured the association between occupations and RID-related presenteeism [24, 29, 31, 40, 42, 52, 53, 60]. In a survey of employees from organisations represented at the Sedgwick County Pandemic Influenza Workgroup, Kansas, healthcare workers were more likely to report previously working with ILI than other workers, including those in education. [31].

Results from a cross-sectional study based on an internet survey of 1,226 Japanese employees [52], showed that company employees are more likely to return to work within 7 days after symptom onset, compared to the self-employed, part-time workers and government workers.

Although the definition of ‘company employee’ is not clearly described in the paper, we would assume that this term refers to full time employment in Japan. In contrast, a study of symptomatic COVID-19 cases in London workplaces, [53] found no differences in workplace attendance after COVID-19 symptom onset between occupational sectors, including office, retail and construction.

In a single study of Canadian healthcare workers, physicians were significantly more likely to work with RTI than medical students and residents, and considered the risk of transmitting infection to others to be the lowest [29]. A study using data from publicly funded healthcare workers in Queensland, Australia, demonstrated that nursing staff and health practitioners had longer sick leave than medical doctors [60]. However, a single centre study of healthcare workers in New York, U.S., [24] reported that physicians and nurses were equally likely to work while symptomatic.

#### Work and employment

Eight studies reported “ lack of cover” as one of the main reasons for presenteeism [19, 26, 28, 41, 44, 49, 51, 61] and one reported an association in healthcare workers [42]. The percentage of participants who chose this as one of the main reasons ranged from 24% to 96%. Four studies reported concerns about “ pay loss” as a main reason [18, 35, 40, 51] and one study reported that attendance bonuses incentivised employees to work while ill [14]. Three studies cited workload and fear of falling behind at work felt as main reasons, categorised as ‘job demand’ [26, 40, 41].

Four studies tested associations between “ paid sick leave” and RID-related presenteeism. A study assessing workers at five U.S. Influenza Vaccine Effectiveness Network sites, who had medically attended ARI or influenza during the 2017–2018 influenza season, [13] reported that workers who had access to paid leave were significantly less likely to work during the first 3 days of illness with an acute respiratory infection (ARI). A cross-sectional study using nationally representative survey data from households across the U.S.[21], reported that employees with paid sick days had a higher probability of staying home for their own and child’s ILI or influenza. Jiang et al [40] reported that healthcare workers from nine Canadian hospitals, who did not receive paid sick leave, were significantly more likely to choose “ can’t afford to stay home” while symptomatic with an ARI. : Hoang Johnson et al [42] identified that access to paid sick leave increases adherence to absenteeism for ILI in healthcare workers at a midwestern academic institution in the U.S.

Four studies tested flexible work or leave policy as a risk factor for RID-related presenteeism [13, 27, 52, 63]. Inflexible work conditions, such as work that does not accommodate home working, appears to be a driver for presenting at work with RID symptoms. Machida et al [52] demonstrated that “ unable to work from home” was a significant factor for going to work within 7 days of symptom onset during the COVID-19 outbreak in 1226 Japanese workers. In addition, a cohort study of employees from three large U.S. employers [27],reported that an employee without a “ work from home” policy is significantly more likely to attend work when ILI symptoms are most severe. Furthermore, a telephone interview survey of a representative sample of Australian adults demonstrated that the intention to comply with home quarantine following exposure to pandemic influenza was much lower for the employed who are unable to work from home, compared to people not in paid employment [63].

#### Social norms and expectations

Eight studies linked presenteeism and social norms and expectations, but only one was outside the healthcare sector [26]. Five studies reported “ avoid burdening colleagues” [17, 35, 36, 39, 61] with 57% to 100% of participants choosing this as a main reason for presenteeism. Five studies investigated “ peer pressure” as a main reason and concerns were expressed as “ avoid or afraid of appearing weak or lazy”, or “ feeling pressure or judgement from colleagues or supervisors” [26, 28, 36, 42, 61]. Rebmann et al [41] reported that “ perceived pressure from colleagues or supervisor” was a significant predictor of presenteeism behaviour among school nurses located in Missouri, U.S.

#### Organisational factors

Three studies cited reasons related to presenteeism culture such as “ had a perception that they were encouraged to work while ill” or “ seeing other colleagues working when similarly unwell” [35, 40, 61]. In addition, Ahmed et al [13] reported that participants were significantly less likely to attend their usual workplace during the first 3 days of ILI if they were discouraged from coming to work when ill. Likewise, a survey of Missouri school nurses indicated that they were more likely to have engaged in presenteeism if their school culture encouraged staff to work while ill [41]. Furthermore, a study of healthcare workers in a tertiary-care healthcare system reported that being directed by management to come into work was a perceived barrier to absenteeism [42].

Reasons related to the perceived threat of disciplinary action or negative repercussions (e.g. reprimand or disapproval) were reported in two studies [36, 42], and a single-centre survey of healthcare workers reported that awareness of outbreak control measures within their facilities appeared to influence their attendance decisions [66].Sociodemographic factors Five studies found significant associations between gender and RID-related presenteeism. Two cross-sectional survey studies of Australian adults, showed females were more likely to report adherence to public health guidance to stay at home following exposure to influenza pandemic, compared to males [62, 63]. Moreover, a study using data from a nationally representative survey of households across the U.S., showed that women are more likely to stay at home for children’s ILI and influenza [21]. Conversely, a UK-based study showed that males were 66% less likely to attend the workplace with COVID-19 symptoms [53] and a study showed that ILI-related presenteeism rates were higher in female healthcare professionals at two inpatient hospital units in the U.S. [20].

Findings from three studies showed that younger age is linked with higher level of presenteeism and lower rate of anticipated compliance. Mossad et al [20] found that presenteeism was significantly higher among those aged below 40 years in a population of healthcare professionals at two inpatient units in the U.S. A study of Australian telephone survey participants [62] reported that those over 55 years were significantly more likely to report anticipated compliance for both the common cold and seasonal influenza. Ablah et al [31] reported that those 30 years old or younger were 2.8 times more likely to report previously working while ill in a sample of employees from organisations represented at the Sedgwick County Pandemic Influenza Workgroup. In comparison, Jain et al [53] reported no association between age and attendance at London workplaces after COVID-19 symptom onset.

#### Health behaviour or perception

“ Not sick enough or considered a minor disease” is the most reported reason for presenteeism, cited by ten studies with this listed as the main reason in 38% and 84% settings [18, 19, 26, 28, 40, 41, 44, 49, 51, 61], and tested as a significant association with non-adherence to absenteeism in clinical healthcare workers [42]. In addition, a survey of Missouri Association of School Nurses members [41] demonstrated that school nurses were significantly more likely to work with symptoms of ILI, if it was considered a mild illness by the participant. Similarly, Jiang et al [40] reported that healthcare workers from nine Canadian hospitals were significantly less likely to work as symptom severity increased.

A single centre qualitative research study on Philadelphian physicians and clinicians highlighted beliefs that it is “ unreasonable” to expect staff will take sick leave until symptoms of RTI resolve as resolution can take several days [38].

#### Vaccination

Three studies tested the association between vaccinated and non-vaccinated groups with inconsistent results. Using data from a national internet panel survey, a U.S. study showed higher prevalence of RID-related presenteeism among the influenza vaccinated healthcare workers [19]. An Iranian study of nursing staff from three teaching hospitals reported no significant difference in the proportion of nurses who had continued to work with an ILI between the vaccinated and unvaccinated groups [46].Another study using internet-based data of employees of the University of Minnesota, found vaccination is associated with statistically significant reductions in days of working while ill among 50-64 years olds[30].

## DISCUSSION

The search and screening process yielded 54 studies. Of these, 94% (n = 51) were observational studies and 56% (n = 30) were from the U.S. Furthermore, 91% (n = 49) of the studies were of low to moderate quality, with a predisposition to response bias and poor sampling strategies. This limits the robustness of the observations, as bias may have occurred to an extent that the results do not adequately represent the target population. In addition, geographical applicability may also be affected by the predominant U.S. bias, as countries have different working practices, health & safety legislation and culture. Despite the limitations, this review identified clear behavioural trends that may impact the effectiveness of workplace COVID-19 control. In this section we discuss possible prevention policies and potential solutions based on the review and within the context of the COVID-19 pandemic.

### Effective isolation policies

In the UK, as in other countries, one of the response measures to control the spread of COVID-19 within the workforce was a requirement to stay at home and self-isolate where an employee or their family has any COVID-19 symptoms [53]. Reported prevalence of presenteeism for those with confirmed RID ranged from 14.1% to 55%, while for suspected RID or potential exposure to RID this ranged from 6.6% to 100%. Similar results were reported in a review of presenteeism relating to all infectious illness, including gastroenteritis [2].

These findings contrast with other included studies where a high percentage (≥94%) of respondents reported intent to adhere with guidance to stay at home, for at least 7 days, during an influenza pandemic [6, 62, 63]. This may indicate that, while workers may acknowledge a workplace or national policy to stay at home with confirmed or suspected RID, their actual behaviour will be adversely affected by other factors, and so additional occupational health policies may be required to enable policies within the workplace. The COVID-19 Rapid Survey of Adherence to Interventions and Responses [CORSAIR] study demonstrated that adherence rates to test, trace and isolate in the UK was low (18.2%), but intention to carry out these behaviours was much higher (around 70%), and this was associated with a number of factors including gender, age, lower socioeconomic grade and working in a key sector [8].

We recommend that a clear isolation from work policy should be supplemented with a range of IPC measures to reduce workplace transmission and could be accompanied with a policy of RID testing of employees in the workplace. As an example, laboratory confirmation of influenza in medical doctors led to an increase in associated sick leave from 16.6% (baseline) to 76.6% [60]. Moreover, 96% of healthcare workers agreed that it is important for ill employees with confirmed influenza infection (positive test result) to stay at home [42].

Our review was unable to distinguish similar symptoms shared between common colds, such as a sore throat, headache, cough and muscle ache, from influenza [67] or other significant RIDs. However, while colds symptoms can include elevated temperature, they rarely include sudden high temperature of 38^°^C or above (fever), so this criterion can be used to reduce influenza transmission. This can be undertaken through the form of temperature checks either at home (self-assessment), randomised workplace checks or as a condition of entry. Workplace based symptom testing and self-isolation polices can form part of an effective methodology to decrease the impact of RID in high-risk occupational settings.

### Perceived expectations and obligations

The Japanese study by Machida et al [52] determined that most of the participants that did not practice strict self-isolation during the COVID-19 pandemic were company employees, when compared to part-time workers, self-employed and government workers. This may indicate that, in this study company workers felt more obligated to go into work with milder symptoms of RID than other workers, possibly due to organisational or peer pressures.

This feeling of obligation was also observed in other studies. The most common reason given by respondents for RID-presenteeism was a feeling of sense of duty or professional obligation, particularly healthcare workers [18, 19, 22, 39, 61]. While other studies [17, 26, 28, 35, 36, 39, 40, 42, 61] listed reasons related to social norms and presenteeism culture such as “ avoid burdening colleagues”, “ peer pressure” and a “ perception that the organisation encouraged working while ill”.

Ahmed et al [13] found that participants who worked in an organisation in which employees were actively discouraged from attending work if they had influenza-like symptoms were significantly less likely to attend their usual workplace compared to those who were not. Clear messaging throughout all levels of the organisation is therefore vital for an effective transmission prevention policy.

### Role of occupational health services

“ Not sick enough” or “ considered a minor disease” was reported as a justification for RID-presenteeism in, mostly, healthcare workers [18, 19, 26, 28, 40, 41, 44, 49, 51, 61]. This indicates that organisations should consider staff education and training regarding the consequences of presenteeism, even when symptoms are minor or employees are asymptomatic, to limit the risk to other employees, visitors, clients or patients when they attend work with RID. Mitchell and Coatsworth [61] noted that there is a role for occupational health services in educating staff regarding the risk of coming to work with an RID. This may be especially relevant where only minor RID symptoms are identified, and to separate common colds from more severe RID.

### Resourcing and contingencies

Presenteeism due to a “ lack of cover” was reported in eight studies, predominantly in healthcare workers [19, 26, 28, 41, 44, 49, 51, 61]. This can be prevented by having contingency plans in place. Miwa et al [44] found that improvements in workplace logistics during times of high RID prevalence, such as providing additional human resources and back-up systems, enabled healthcare workers to feel they could take sick leave when necessary.

### Pay and bonus schemes

Four studies reported “ pay loss” as a reason for presenteeism [18, 35, 40, 51]. Babcock et al [35] suggested that, where organisations combine paid vacation and sick days, a worker might decide to go to work with RID rather than claim it as a sick day and, thus, lose a vacation day. Their recommendation is that this policy is avoided as it created “ *a perverse incentive for presenteeism*”.

Attendance bonuses may also incentivise employees to work while ill [14], with the potential for bonuses to be missed regardless of cause [35]. This may, however, have direct and indirect effects on organisations due to health-related productivity loss which can lead to economic costs. For example, Letvak et al [68] found that nurse presenteeism in U.S. hospitals raised health care costs, with estimated costs of about $2 billion dollars annually from increased patient falls, medication errors and lower quality-of-care scores. Hence, organisations should consider unintended consequences of RID and other forms of presenteeism, such as economic cost and employee health and wellbeing, before implementing attendance bonus schemes, respectively, due to presenteeism.

Paid sick leave was significantly negatively associated with lower RID-presenteeism in three studies [13, 21, 40]. While annual leave and paid sick leave is statutory in many European countries, as more workers become self-employed contractors to online platforms such as Amazon and Uber [69], access to employee benefits may decline. Most self-employed workers do not receive paid sick leave and have less collective bargaining power against platform companies [70, 71]. Although companies such as Just Eat and Deliveroo offered some financial support, i.e., for a maximum of 14 days with certain eligibility criteria, for their takeaway couriers who were required to self-isolate over the COVID-19 pandemic [72, 73].

### Employee demographics

Gender was significantly associated with RID-presenteeism. Women had a higher prevalence of RID presenteeism [20, 53] but conversely demonstrated a higher probability of staying home for a child’s RID [21]. Studies show that economic slowdowns disproportionately affect women. According to analysis by McKinsey and Company, women represented 39% of the global workforce but accounted for 54% of job losses in 2020 [74]. Mothers, particularly lone mothers, were more likely to work for sectors that had been shut down by the UK government during the COVID-19 pandemic [75]. This may account for the higher incidence of COVID-19 presenteeism in women in the UK, identified by Jain et al [53]. Conversely, women not working in lockdown sectors are twice as likely as men to be key workers, and over four times as likely to work for the health and social care sector [75]. Mothers of younger children in the U.S. reduced their work hours four to five times more than fathers during the COVID-19 pandemic [76]. Therefore, the ability for women to do paid work may be adversely impacted by the pressures from childcare even if their jobs remain active. Organisations should engage with employees when reviewing contingency plans and policies to ensure they do not generate unequal impacts.

### Vaccination Policy

This review identified inconsistent results for the association between receiving influenza vaccination and presenteeism. Askarian et al [46] found no significant difference in continuing to work with ILI symptoms between vaccinated and unvaccinated nurses. Chiu et al [19] reported that the uptake of influenza vaccine was associated with ILI-related presenteeism and that vaccination may reduce symptomatic healthcare workers’ perceived risk of having influenza and onwards transmission. Conversely, Nichol et al [30] reported that influenza vaccination uptake is associated with a reduction in days of working with ILI.

Although influenza vaccinations interrupt viral transmission and reduce illness, there is an annual variation in effectiveness. For example, studies have found vaccine effectiveness to be reduced in those who received repeated prior influenza vaccinations [77, 78]. There is the potential for an adverse relationship to develop between vaccination uptake and presenteeism, with vaccine recipients willing to work while ill, because they have been vaccinated, yet unaware of the limitations on the effectiveness.

Therefore, we recommend that where an organisation has a vaccination policy consideration, or takes advantage of a national programme, employees should still be advised on the steps to take if COVID-19 symptoms are reported with regards to attending the workplace.

## CONCLUSION

Recent research on RID-related workplace presenteeism, including COVID-19, has provided further understanding of the associated risk factors. While there is a strong intention among workers to adhere to non-pharmaceutical interventions, such as requirements to stay at home with an RID, studies still showed high levels of presenteeism. Factors linked to presenteeism include organisational culture, such as organisational or peer pressure, ineffective resource planning preventing workers from taking time off, inadequate sick leave / sick pay, attendance-based bonus schemes, lack of occupational health services and gender inequality. The inter-relationship of the factors associated with presenteeism means effective non-pharmaceutical interventions require a comprehensive review of related supporting workforce and organisational policies.

There was insufficient research on the potential impact of onsite RID testing as a preventative factor for presenteeism, while impact of vaccine was inconclusive. With a bias towards health and social care workers, future research should focus on the role of respiratory infection testing and vaccination as intervention strategies for vulnerable and precarious occupational groups, including self-employed and gig workers.

## Data Availability

All relevant data are either included in this article or added to the supplementary information.

## Additional files

Additional file 1: Search strategy used in MEDLINE. Word document (.docx)

Additional file 2: Study characteristics. Word document (.docx)

Additional file 3: Quality assessment of the included studies. Word document (.docx)

Additional file 4: Presenteeism measurements for the included studies. Word document (.docx)

Additional file 5: Reported reasons for presenteeism. Word document (.docx)

Additional file 6: Risk factors for presenteeism. Word document (docx)

### Abbreviations

ARI: Acute respiratory infection
CASP: Critical Appraisal Skills Programme
CNKI: China Knowledge Resource Integrated Database
ILI: Influenza like illness
IPC: Infection prevention control
NOS: Newcastle-Ottawa Scale
PRISMA: Preferred Reporting Items for Systematic Reviews and Meta-Analyses
RID: Respiratory infectious disease
RTI: respiratory tract infection
SARS-CoV-2: Severe acute respiratory syndrome coronavirus 2

## Declarations

### Ethics approval and consent to participate

Not applicable

### Consent for publication

Not applicable

### Availability of data and materials

The datasets used and/or analysed during the current study available from the corresponding author on reasonable request.

## Ethics approval

Ethical approval was not required because no sensitive or confidential information was collected from participants.

## Authors’ contributions

SD and HW made substantial contributions to the design of the study and carried out the full search. SD, HW and YH screened the literature for inclusion and exclusion, with support from HC and MvT. SD and HW performed the analysis and interpretation of the data. SD and HW drafted the first version. Substantial revisions were provided by HC, DD, IH, MR, AV, CW and MvT. All authors read and approved the final version of the manuscript.

## Acknowledgements

Not applicable

## References

1. WHO: Modes of transmission of virus causing COVID-19: implications for IPC precaution recommendations: scientific brief, 29 March 2020. In. Geneva: World Health Organization; 2020.

2. Webster RK, Liu R, Karimullina K, Hall I, Amlôt R, Rubin GJ: A systematic review of infectious illness Presenteeism: prevalence, reasons and risk factors. BMC Public Health 2019, 19(1):799.

3. Ruhle SA, Breitsohl H, Aboagye E, Baba V, Biron C, Correia Leal C, Dietz C, Ferreira AI, Gerich J, Johns G et al: “ To work, or not to work, that is the question” – Recent trends and avenues for research on presenteeism. European Journal of Work and Organizational Psychology 2020, 29(3):344–363.

4. Anderson NJ, Bonauto DK, Fan ZJ, Spector JT: Distribution of Influenza-Like Illness (ILI) by Occupation in Washington State, September 2009–August 2010. PLOS ONE 2012, 7(11):e48806.

5. Ladhani SN, Chow JY, Janarthanan R, Fok J, Crawley-Boevey E, Vusirikala A, Fernandez E, Perez MS, Tang S, Dun-Campbell K et al: Increased risk of SARS-CoV-2 infection in staff working across different care homes enchanced COVID-19 outbreak investigations in London care homes. Journal of Infection 2020, 81(4): 621–624.

6. Blendon RJ, Koonin LM, Benson JM, Cetron MS, Pollard WE, Mitchell EW, Weldon KJ, Herrmann MJ: Public response to community mitigation measures for pandemic influenza. Emerg Infect Dis 2008, 14(5):778–786.

7. Blake KD, Blendon RJ, Viswanath K: Employment and compliance with pandemic influenza mitigation recommendations. Emerging Infectious Diseases 2010, 16(2):212–218.

8. Smith LE, Potts HWW, Amllt R, Fear NT, Michie S, Rubin GJ: Adherence to the test, trace and isolate system: results from a time series of 21 nationally representative surveys in the UK (the COVID-19 Rapid Survey of Adherence to Interventions and Responses [CORSAIR] study). medRxiv 2020:2020.2009.2015.20191957.

9. Stevens A, Garritty C, Hersi M, Moher D: Developing PRISMA-RR, a reporting guideline for rapid reviews of primary studies (Protocol). In.: Oxford, UK: EQUATOR Network; 2018.

10. Johns G: Presenteeism in the workplace: A review and research agenda. Journal of Organizational Behavior 2010, 31(4):519–542.

11. Houghton C, Meskell P, Delaney H, Smalle M, Glenton C, Booth A, Chan XHS, Devane D, Biesty LM: Barriers and facilitators to healthcare workers’ adherence with infection prevention and control (IPC) guidelines for respiratory infectious diseases: a rapid qualitative evidence synthesis. Cochrane Database Syst Rev 2020, 4(4):Cd013582.

12. Herzog R, Álvarez-Pasquin MJ, Díaz C, Del Barrio JL, Estrada JM, Gil Á: Are healthcare workers’ intentions to vaccinate related to their knowledge, beliefs and attitudes? a systematic review. BMC Public Health 2013, 13(1):154.

13. Ahmed F, Kim S, Nowalk MP, King JP, VanWormer JJ, Gaglani M, Zimmerman RK, Bear T, Jackson ML, Jackson LA et al: Paid leave and access to telework as work attendance determinants during acute respiratory illness, United States, 2017-2018. Emerging Infectious Diseases 2020, 26(1):26–33.

14. Dyal JW, Grant MP, Broadwater K, Bjork A, Waltenburg MA, Gibbins JD, Hale C, Silver M, Fischer M, Steinberg J et al: COVID-19 Among Workers in Meat and Poultry Processing Facilities - 19 States, April 2020. MMWR Morbidity and mortality weekly report 2020, 69(18).

15. Kobayashi M, Lyman MM, Francois Watkins LK, Toews KA, Bullard L, Radcliffe RA, Beall B, Langley G, Beneden CV, Stone ND: A Cluster of Group A Streptococcal Infections in a Skilled Nursing Facility-the Potential Role of Healthcare Worker Presenteeism. J Am Geriatr Soc 2016, 64(12):e279–e284.

16. Magill SS, Black SR, Wise ME, Kallen AJ, Lee SJ, Gardner T, Husain F, Srinivasan A, Gerber SI, Jhung M: Investigation of an outbreak of 2009 pandemic influenza A virus (H1N1) infections among healthcare personnel in a Chicago hospital. Infect Control Hosp Epidemiol 2011, 32(6):611–615.

17. Cowman K, Mittal J, Weston G, Harris E, Shapiro L, Schlair S, Park S, Nori P: Understanding drivers of influenza-like illness presenteeism within training programs: A survey of trainees and their program directors. American Journal of Infection Control 2019, 47(8):895–901.

18. Wilson KE, Wood SM, Schaecher KE, Cromwell KB, Godich J, Knapp MH, Sklar MJ, Ewing D, Raviprakash K, Defang G et al: Nosocomial outbreak of influenza A H3N2 in an inpatient oncology unit related to health care workers presenting to work while ill. American Journal of Infection Control 2019, 47(6):683–687.

19. Chiu S, Black CL, Yue X, Greby SM, Laney AS, Campbell AP, de Perio MA: Working with influenza-like illness: Presenteeism among US health care personnel during the 2014-2015 influenza season. American journal of infection control 2017, 45(11):1254–1258.

20. Mossad SB, Deshpande A, Schramm S, Liu X, Rothberg MB: Working Despite Having Influenza-Like Illness: Results of An Anonymous Survey of Healthcare Providers Who Care for Transplant Recipients. Infection Control and Hospital Epidemiology 2017, 38(8):966–969.

21. Piper K, Youk A, James AE, 3rd, Kumar S: Paid sick days and stay-at-home behavior for influenza. PLoS One 2017, 12(2):e0170698.

22. de Perio MA, Wiegand DM, Brueck SE: Influenza-like illness and presenteeism among school employees. American Journal of Infection Control 2014, 42(4):450–452.

23. Norton DM, Brown LG, Frick R, Carpenter LR, Green AL, Tobin-D’Angelo M, Reimann DW, Blade H, Nicholas DC, Egan JS et al: Managerial practices regarding workers working while ill. J Food Prot 2015, 78(1):187–195.

24. Bhadelia N, Sont R, McCarthy JW, Vorenkamp J, Jia HM, Saiman L, Furuya EY: Impact of the 2009 Influenza A (H1N1) Pandemic on Healthcare Workers at a Tertiary Care Center in New York City. Infection Control and Hospital Epidemiology 2013, 34(8):825–831.

25. Esbenshade JC, Edwards KM, Esbenshade AJ, Rodriguez VE, Talbot HK, Joseph MF, Nwosu SK, Chappell JD, Gern JE, Williams JV et al: Respiratory virus shedding in a cohort of on-duty healthcare workers undergoing prospective surveillance. Infection Control and Hospital Epidemiology 2013, 34(4):373–378.

26. Rebmann T, Wang J, Swick Z, Reddick D, Minden-Birkenmaier C: Health care versus non-health care businesses’ experiences during the 2009 H1N1 pandemic: financial impact, vaccination policies, and control measures implemented. Am J Infect Control 2013, 41(6):e49–54.

27. Rousculp MD, Johnston SS, Palmer LA, Chu B-C, Mahadevia PJ, Nichol KL: Attending work while sick: Implication of flexible sick leave policies. Journal of occupational and environmental medicine (Online) 2010, 52(10):1009–1013.

28. May L, Katz R, Johnston L, Sanza M, Petinaux B: Assessing physicians’ in training attitudes and behaviors during the 2009 H1N1 influenza season: a cross-sectional survey of medical students and residents in an urban academic setting. Influenza and Other Respiratory Viruses 2010, 4(5):267–275.

29. Gudgeon P, Wells DA, Baerlocher MO, Detsky AS: Do you come to work with a respiratory tract infection? Occup Environ Med 2009, 66(6):424.

30. Nichol KL, D’Heilly SJ, Greenberg ME, Ehlinger E: Burden of influenza-like illness and effectiveness of influenza vaccination among working adults aged 50-64 years. Clin Infect Dis 2009, 48(3):292–298.

31. Ablah E, Konda K, Tinius A, Long R, Vermie G, Burbach C: Influenza vaccine coverage and presenteeism in Sedgwick County, Kansas. American Journal of Infection Control 2008, 36(8):588–591.

32. LaVela S, Goldstein B, Smith B, Weaver FM: Working with symptoms of a respiratory infection: Staff who care for high-risk individuals. American Journal of Infection Control 2007, 35(7):448–454.

33. de Perio MA, Brueck SE, Mueller CA, Milne CK, Rubin MA, Gundlapalli AV, Mayer J: Evaluation of 2009 pandemic influenza A (H1N1) exposures and illness among physicians in training. American Journal of Infection Control 2012, 40(7):617–621.

34. Palmer LA, Rousculp MD, Johnston SS, Mahadevia PJ, Nichol KL: Effect of influenza-like illness and other wintertime respiratory illnesses on worker productivity: The child and household influenza-illness and employee function (CHIEF) study. Vaccine 2010, 28(31):5049–5056.

35. Babcock HM, Beekmann SE, Pillai SK, Santibanez S, Lee L, Kuhar DT, Campbell AP, Patel A, Polgreen PM: Reported variability in healthcare facility policies regarding healthcare personnel working while experiencing influenza-like illnesses: An emerging infections network survey. Infection Control and Hospital Epidemiology 2020, 41(1):80–85.

36. Kaldjian LC, Shinkunas LA, Reisinger HS, Polacco MA, Perencevich EN: Attitudes about sickness presenteeism in medical training: is there a hidden curriculum? Antimicrobial Resistance and Infection Control 2019, 8:149.

37. O’Neil CA, Kim L, Prill MM, Stone ND, Garg S, Talbot HK, Babcock HM: Preventing Respiratory Viral Transmission in Long-Term Care: Knowledge, Attitudes, and Practices of Healthcare Personnel. Infection Control and Hospital Epidemiology 2017, 38(12):1449–1456.

38. Szymczak JE, Smathers S, Hoegg C, Klieger S, Coffin SE, Sammons JS: Reasons Why Physicians and Advanced Practice Clinicians Work While Sick: A Mixed-Methods Analysis. JAMA Pediatr 2015, 169(9):815–821.

39. Jena AB, Meltzer DO, Press VG, Arora VM: Why physicians work when sick. Arch Intern Med 2012, 172(14):1107–1108.

40. Jiang L, McGeer A, McNeil S, Katz K, Loeb M, Muller MP, Simor A, Powis J, Kohler P, Di Bella JM et al: Which healthcare workers work with acute respiratory illness? Evidence from Canadian acute-care hospitals during 4 influenza seasons: 2010-2011 to 2013-2014. Infection Control and Hospital Epidemiology 2019, 40(8):889–896.

41. Rebmann T, Turner JA, Kunerth AK: Presenteeism Attitudes and Behavior Among Missouri Kindergarten to Twelfth Grade (K-12) School Nurses. The Journal of school nursing : the official publication of the National Association of School Nurses 2016, 32(6):407–415.

42. Hoang Johnson D, Osman F, Bean J, Stevens L, Shirley D, Keating JA, Johnson S, Safdar N: Barriers and facilitators to influenza-like illness absenteeism among healthcare workers in a tertiary-care healthcare system, 2017-2018 influenza season. Infect Control Hosp Epidemiol 2021:1–8.

43. Podewils LJ, Burket TL, Mettenbrink C, Steiner A, Seidel A, Scott K, Cervantes L, Hasnain-Wynia R: Disproportionate Incidence of COVID-19 Infection, Hospitalizations, and Deaths Among Persons Identifying as Hispanic or Latino - Denver, Colorado March-October 2020. MMWR Morb Mortal Wkly Rep 2020, 69(48):1812–1816.

44. Miwa T, Tagashira Y, Uenoyama Y, Honda H: Healthcare workers’ presenteeism and chemoprophylaxis against nosocomial influenza in patients hospitalized during the 2018-2019 season. J Hosp Infect 2020, 106(2):399–400.

45. Yu J, Ren X, Ye C, Tian K, Feng L, Song Y, Cowling BJ, Li Z: Influenza Vaccination Coverage among Registered Nurses in China during 2017-2018: An Internet Panel Survey. Vaccines (Basel) 2019, 7(4):134.

46. Askarian M, Khazaeipour Z, McLaws ML: Facilitators for influenza vaccination uptake in nurses at the Shiraz University of Medical Sciences. Public Health 2011, 125(8):512–517.

47. Fernando M, Caputi P, Ashbury F: Impact on Employee Productivity From Presenteeism and Absenteeism: Evidence From a Multinational Firm in Sri Lanka. Journal of Occupational & Environmental Medicine 2017, 59(7):691–696.

48. Li Y, Edwards J, Huang B, Shen C, Cai C, Wang Y, Zhang G, Robertson I: Risk of zoonotic transmission of swine influenza at the human-pig interface in Guangdong Province, China. Zoonoses Public Health 2020, 67(6):607–616.

49. JiF 季, Sun J, Shang S 尚: 呼吸内科医 人 医院呼吸道感染的因 素分析及 策 (Analysis of factors and measures for hospital respiratory tract infection of medical staff in department of respiratory medicine). •••理•志 (Journal of Qilu Nursing) 2008, 14(23):122–123.

50. Rao F, Huang Z 黄, Zhang Q, Zhong X, Liu Y 刘, Zhang M, Yuan R 袁, Lu Z : 莞市中小学生、工人和居民甲型 H1N1 流感防治知信行 况 (Cross-sectional Study on Knowledge,Attitude and Behavior of Influenza A(H1N1) Among Students,Workers and Residents in Dongguan). • 用• 防医学 (Practical Preventive Medicine) 2010, 17(11):2184–2187.

51. Wang P 王, Dong W 董薇, Yang S: 广州市三甲医院 士流感 病例流行病学研究(An Epidemiological Study on Influenza-like Cases among Nurses in Guangzhou’s Tier 3 Hospitals). 吉林医学 (Jilin Medical Journal) 2012, 33(21):4579–4580.

52. Machida M, Nakamura I, Saito R, Nakaya T, Hanibuchi T, Takamiya T, Odagiri Y, Fukushima N, Kikuchi H, Amagasa S et al: The actual implementation status of self-isolation among Japanese workers during the COVID-19 outbreak. Tropical medicine and health 2020, 48(1):63–63.

53. Jain V, Waghorn M, Thorn-Heathcock R, Lamb P, Bell A, Addiman S: Attendance at London workplaces after symptom onset: a retrospective cohort study of staff members with confirmed COVID-19. Journal of Public Health 2021, 43(2):236–242.

54. Mikos M, Juszczyk G, Czerw A, Strzepek Banas T, Cipora E, Deptala A, Badowska-Kozakiewicz A: Refusal to Take Sick Leave after Being Diagnosed with a Communicable Disease as an Estimate of the Phenomenon of Presenteeism in Poland. Medical Principles and Practice 2020, 29(2):134–141.

55. Juszczyk G, Czerw A, Augustynowicz A, Banas T, Mikos M, Religioni U, Deptala A: Refusal to take a sick leave as an estimate of the phenomenon of presenteeism in Poland. Oncotarget 2018, 9(46):28176–28184.

56. Martinez LF, Ferreira AI: Sick at work: presenteeism among nurses in a Portuguese public hospital. Stress Health 2012, 28(4):297–304.

57. Iverson D, Lewis KL, Caputi P, Knospe S: The cumulative impact and associated costs of multiple health conditions on employee productivity. J Occup Environ Med 2010, 52(12):1206–1211.

58. Meilicke G, Riedmann K, Biederbick W, Muller U, Wierer T, Bartels C: Hygiene perception changes during the influenza A H1N1 pandemic in Germany: incorporating the results of two cross-sectional telephone surveys 2008-2009. BMC public health 2013, 13:959.

59. Kuster SP, Böni J, Kouyos RD, Huber M, Schmutz S, Shah C, Bischoff-Ferrari HA, Distler O, Battegay E, Giovanoli P et al: Absenteeism and presenteeism in healthcare workers due to respiratory illness. Infect Control Hosp Epidemiol 2020:1–6.

60. Imai C, Hall L, Lambert SB, Merollini KMD: Presenteeism among health care workers with laboratory-confirmed influenza infection: A retrospective cohort study in Queensland, Australia. American Journal of Infection Control 2020, 48(4):355–360.

61. Mitchell L, Coatsworth N: Sick leave accessibility in junior doctors at an Australian health service. Infection, Disease & Health 2020, 26(1):3–10.

62. Brown LH, Aitken P, Leggat PA, Speare R: Self-reported anticipated compliance with physician advice to stay home during pandemic (H1N1) 2009: results from the 2009 Queensland Social Survey. BMC Public Health 2010, 10:138.

63. Eastwood K, Durrheim D, Francis JL, d’Espaignet ET, Duncan S, Islam F, Speare R: Knowledge about pandemic influenza and compliance with containment measures among Australians. Bull World Health Organ 2009, 87(8):588–594.

64. Tartari E, Saris K, Kenters N, Marimuthu K, Widmer A, Collignon P, Cheng VCC, Wong SC, Gottlieb T, Tambyah PA et al: Not sick enough to worry? “ Influenza-like” symptoms and work-related behavior among healthcare workers and other professionals: Results of a global survey. PLoS One 2020, 15(5):e0232168.

65. McGregor A, Ashbury F, Caputi P, Iverson D: A preliminary investigation of health and work-environment factors on presenteeism in the workplace. Journal of Occupational and Environmental Medicine 2018, 60(12):E671–E678.

66. LaVela S, Goldstein B, Smith B, Weaver FM: Working with symptoms of a respiratory infection: staff who care for high-risk individuals. Am J Infect Control 2007, 35(7):448–454.

67. NHS: Health A to Z. https://www.nhs.uk/conditions/. Accessed 29 March 2021.

68. Letvak SA, Ruhm CJ, Gupta SN: Nurses’ presenteeism and its effects on self-reported quality of care and costs. Am J Nurs 2012, 112(2):30–38; quiz 48, 39.

69. Tran M, Sokas RK: The Gig Economy and Contingent Work: An Occupational Health Assessment. J Occup Environ Med 2017, 59(4):e63–e66.

70. Peckham T, Fujishiro K, Hajat A, Flaherty BP, Seixas N: Evaluating Employment Quality as a Determinant of Health in a Changing Labor Market. Rsf 2019, 5(4):258–281.

71. Rönnblad T, Grönholm E, Jonsson J, Koranyi I, Orellana C, Kreshpaj B, Chen L, Stockfelt L, Bodin T: Precarious employment and mental health: a systematic review and meta-analysis of longitudinal studies. Scand J Work Environ Health 2019, 45(5):429–443.

72. Deliveroo: Latest COVID-19 Updates. https://riders.deliveroo.co.uk/en/news/latest-covid-19-updates. Accessed 29 March 2021.

73. JustEat: Just Eat UK’s financial support for couriers on the Just Eat Network. https://www.just-eat.co.uk/info/covid-19/courier-financial-support. Accessed 29 March 2021.

74. McKinseyandCompany: COVID-19 and Gender Equality: Countering the Regressive Effects. https://www.mckinsey.com/featured-insights/future-of-work/covid-19-and-gender-equality-countering-the-regressive-effects. Accessed 29 March 2021.

75. Blundell R, Costa Dias M, Joyce R, Xu X: COVID-19 and Inequalities. Fiscal studies 2020, 41(2):291–319.

76. Collins C, Landivar LC, Ruppanner Scarborough WJ: COVID-19 and the Gender Gap in Work Hours. Gender, Work & Organization 2021, 28(S1):101–112.

77. Skowronski DM, Chambers C, Sabaiduc S, De Serres G, Winter AL, Dickinson JA, Krajden M, Gubbay JB, Drews SJ, Martineau C et al: A Perfect Storm: Impact of Genomic Variation and Serial Vaccination on Low Influenza Vaccine Effectiveness During the 2014-2015 Season. Clin Infect Dis 2016, 63(1):21–32.

78. Ramsay LC, Buchan SA, Stirling RG, Cowling BJ, Feng S, Kwong JC, Warshawsky BF: The impact of repeated vaccination on influenza vaccine effectiveness: a systematic review and meta-analysis. BMC Med 2019, 17(1):9.

