## Additional file 1 for "Risk factors associated with respiratory infectious disease-related presenteeism: a rapid review"

*Additional file 1: Search strategy used in MEDLINE*

1. exp presenteeism/ or presenteeism.mp.

2. ((going to work or working) adj3 (ill* or sick*) ).mp.

3. ((suspected adj2 symptom*) or (like adj2 symptom*) or (mild* adj2 symptom*) or subclinical or probat* or prodrom*).mp.

4. ((going to work or working) adj3 ((suspected adj2 symptom*) or (like adj2 symptom*) or (mild* adj2 symptom*) or subclinical or probat* orprodrom*)).mp.

5. (against or "not adhere to" or violat* or non-complian* or "not comply*" or "lack of compliance").mp.

6. (guideline or guidance or protocol).mp.

7. (isolat* or quarantine or social distanc* or lockdown or lock-down).mp.

8. 5 and ((guideline or guidance or protocol) adj3 (isolat* or quarantine or social distanc* or lockdown or lock-down)).mp.

9. ((expos* or "contact with") adj3 (confirmed or diagnosed or suspected)).mp.

10. (going to work or working).mp. and 9

11. 1 or 2 or 4 or 8 or 10

12. (covid or coronavirus or ncov or sars or sars-cov* or mers or flu or influenza or influenza-like or respiratory infectious disease* or tuberculosis or TB or respiratory tract infection or RTI).mp.

13. 11 and 12

14. remove duplicates from 13
