## Additional file 2 for "Risk factors associated with respiratory infectious disease-related presenteeism: a rapid review"

Additional file 2: Study characteristics

| *Additional file 2: Study characteristics* | | | | | | |
| --- | --- | --- | --- | --- | --- | --- |
|  | Study | Country | Study type | Population type | Sample number; age; % female | RID studied |
| Working while infected (confirmed) | Jain et al., 2021 [53] | UK | Retrospective cohort | Workers on the London Coronavirus Response Centre database that records COVID-19 cases | 130; 18- >56; 36.2 | COVID-19 |
|  | Imai et al., 2020 [60] | Australia | Retrospective cohort | HCWs | 1323; nr; nr | Influenza |
|  | Mikos et al., 2020 [54] | Poland | Cross sectional | Patients who were professionally active, either employed or running their own business | 2529; M = 36.4; 56.9 | Influenza |
|  | Juszczyk et al., 2018 [55] | Poland | Cross sectional | Patients who were professionally active, either employed or running their own business | 550360; M = 35.8; 61.9 | ICD-10 codes J0-J4 |
|  | Kobayashi et al., 2017 [15] | USA | Cross sectional | Skilled nursing facility residents and staff | 162; nr; nr | Group A streptococcus |
|  | Magill et al., 2012 [16] | USA | Cross sectional | HCPs | 20; MED = 34; 65 | pH1N1 |
| Working with symptoms (suspected or subclinical) | Hoang Johnson et al., 2021 [42] | USA | Cross sectional | HCWs (clinical and non-clinical) | 2391; nr; 83.5 | ILI |
|  | Ahmed at al., 2020 [13] | USA | Cross sectional | Workers who had medically attended for an ARI with cough within 7 days of illness onset at outpatient facilities affiliated with the IVEN. | 1374; MED = 42; 64 | ARI and confirmed influenza |
|  | Kuster et al., 2021 [59] | Switzerland | Prospective cohort | HCWS in the Acute Care Hospital Setting | 152; 31; 81.6 | Influenza |
|  | Machida et al., 2020 [52] | Japan | Cross sectional | Workers living in seven prefectures | 1226; M = 46.3; 40 | COVID-19 |
|  | Mitchell and Coatsworth, 2020  [61] | Australia | Qualitative | Junior doctors | 54; nr; nr | ARI |
|  | Miwa et al., 2020 [44] | Japan | Cross sectional | HCWs | 90; nr; nr | ILI and confirmed influenza |
|  | Podewils et al., 2020 [43] | USA | Cross sectional | adults aged ≥18 years living in the Denver community | 10163; 18 - >65; 52.3 | COVID-19 |
|  | Cowman et al., 2019 [17] | USA | Cross sectional | Trainees and program leaders in healthcare | 197; 21- >31; 50 | ILI |
|  | Jiang et al., 2019 [40] | Canada | Prospective cohort | HCWs | 2093; MED=41.5; 85.2 | ARI |
|  | Wilson et al., 2019 [18] | USA | Cross sectional | HCWs who had confirmed or probable influenza from November 19-27^th^ 2017 | 14; nr; nr | Influenza A H3N2 |
|  | Yu et al., 2019 [45] | China | Cross sectional | Nurses | 4153; <25- >35; 97.0 | ILI |
|  | McGregor et al., 2018 [64] | nr | Cross sectional | LinkedIn members | 229; <30- >50; 64 | Cold and flu |
|  | Chiu et al., 2017 [19] | USA | Cross sectional | HCPs | 414; 18- >50; nr | ILI |
|  | Mossad et al., 2017 [20] | USA | Cross sectional | HCPs | 286; MED = 35; 72.0 | ILI |
|  | Piper at el., 2017 [21] | USA | Cross sectional | US employees | 12044; M=40.4; 50.9 | ILI and influenza |
|  | Rebmann et al., 2016 [41] | USA | Cross sectional | School nurses | 133; <40- >61; 99.2 | ILI |
|  | Norton et al., 2015 [23] | USA | Cross sectional | Restaurant workers and managers | 426; nr; nr | cold and flu |
|  | De Perio et al., 2014 [22] | USA | Cross sectional | School employees including teachers, admin, bus drivers, food workers and maintenance workers | 412; MED = 46; 82.0 | ILI |
|  | Bhadelia et al., 2013 [24] | USA | Cross sectional | HCWs | 352; 21-68; 75.0 | ILI and confirmed influenza |
|  | Esbenshade et al., 2013 [25] | USA | Cross sectional | HCWs | 170; MED = 31; 81.8 | ILI |
|  | Rebmann et al., 2013 [26] | USA | Cross sectional | HRPs | 471; 21- >61: 7.2%; 83.9 | H1N1 influenza |
|  | Wang et al., 2012 [51] | China | Cross sectional | Nurses | 236; M = 28.5; 94.9 | ILI |
|  | Martinez and Ferreira, 2012 [56] | Portugal | Cross sectional | Nurses | 296; M = 35.7; 72.3 | Breath infections |
|  | Jena et al., 2012 [39] | USA | Cross sectional | Resident physicians | 150; nr; nr | Flu-like symptoms |
|  | Askarian et al., 2011 [46] | Iran | Cross sectional | Nursing staff | 167; MED = 29.8; 91.6 | ILI |
|  | Rousculp et al., 2010 [27] | USA | Prospective cohort | Employees from three US employers | 793; M = 40.7; 35.6 | ILI |
|  | Iverson et al., 2010 [57] | Germany | Cross sectional | Employees of a large multinational company | 667; <30 - >50; 55.9 | Cold and influenza |
|  | May et al., 2010 [28] | USA | Cross sectional | Medical students and residents at an urban institution | 67; M = 31; 50 | ILI |
|  | Gudgeon et al., 2009 [29] | Canada | Cross sectional | Staff and students from the University of Toronto | 149 medical students, 317 residents, 202 physicians; nr; nr | RTI |
|  | Nichol et al., 2009 [30] | USA | Prospective cohort | Employees of the University of Minnesota | 497; 50-64; 77.5 | ILI |
|  | Ji et al., 2008 [49] | China | Cross sectional | Respiratory internal medicine healthcare workers | 405; nr; nr | Upper respiratory tract infection |
|  | Ablah et al., 2008 [31] | USA | Cross sectional | Workers from companies represented by the Sedgwick County Pandemic Influenza Workgroup | 1485; <30- >60; 72 | ILI |
|  | Fernando et al., 2008 [47] | Sri Lanka | Cross sectional | Staff at the headquarters of a multinational firm | 150; <30- >50; 22.5 | Cold and influenza |
|  | LaVela et al., 2007 [32] | USA | Cross sectional | HCWs at spinal cord injury centres | 753; <25- >65; 73 | Respiratory infection |
| Going to work with history of exposure to an RID | De Perio et al., 2012 [33] | USA | Cross sectional | Trainee physicians | 88; MED = 30; 35 | pH1N1 or ILI |
|  | Palmer et al., 2010 [34] | USA | Prospective Cohort | Employees from three large U.S. companies, who had at least one child at home | 2013; M = 41.7; 31.3 | ARI |
| Intention to attend work with an RID | Babcock et al., 2020 [35] | USA | Qualitative | Physicians | 51; nr; nr | ILI |
|  | Dyal et al., 2020 [14] | USA | Qualitative | Workers from 115 meat and poultry facilities | 130578; nr; nr | COVID-19 |
|  | Li et al., 2020 [48] | China | Cross sectional | Pig farmers, traders and trade workers | 190; nr; nr | Swine influenza |
|  | Tartari et al., 2020 [64] | Worldwide | Cross sectional | HCWs and other professionals | 533; 18- >50; 70.5 | ILI |
|  | Kaldjian et al., 2019 [36] | USA | Cross sectional | Medical students and physicians | 127; nr; nr | Cold and flu |
|  | O’Neil et al., 2017 [37] | USA | Cross sectional | Healthcare personnel at a long-term care facility | 73; nr; 67.1 | Respiratory infection |
|  | Szymczak et al., 2015 [38] | USA | Cross sectional | Attending physicians and APCs | 280 (physicians) and 256 (APCs); nr; nr | Respiratory symptoms |
|  | Meilicke et al., 2013 [58] | Germany | Cross sectional | Workers | 2006; 18- >60; 61.1 | Cold symptoms |
|  | Rao et al., 2010 [50] | China | Cross sectional | Students, workers and residents | 1505 (workers 390); M = 18; 52.2 | Influenza |
| Adherence to guidance to stay at home from work with an RID | Brown et al., 2010 [62] | Australia | Cross sectional | Queensland households (subsets of employed and unemployed) | 1292; 18- >55; 49.8 | H1N1 pandemic influenza, avian influenza, seasonal influenza, and common cold |
|  | Eastwood et al., 2009 [63] | Australia | Cross sectional | Adult population (subsets of self-employed, wage earners and unemployed) | 1166; 18- >65; 61.6 | Pandemic influenza |
|  | Blendon et al., 2008 [6] | USA | Cross sectional | Adult population with a subset of employed respondents | 1697 (1101 were employed respondents); 18- >50; 52 | Pandemic flu |

Abbreviations: *ARI* acute respiratory infection; *ILI* influenza-like illness; *HCPs* healthcare professionals; *HCWs* healthcare workers; *HRPs* human resource professionals; *IVEN* Influenza Vaccine Effectiveness Network; *M* mean; *MED* median; *nr* not reported; *RID* respiratory infection disease.
