## Additional file 3 for "Risk factors associated with respiratory infectious disease-related presenteeism: a rapid review"

Additional file 3: Quality assessment of the included studies

| Additional file 3: Quality assessment of the included studies | | | |
| --- | --- | --- | --- |
|  | Reference | Quality assessment tool | Quality |
| Working while infected (confirmed) | Jain et al., 2021 [53] | NOS | High |
|  | Imai et al., 2020 [60] | NOS | High |
|  | Mikos et al., 2020 [54] | Modified NOS | Moderate |
|  | Juszczyk et al., 2018 [55] | Modified NOS | Moderate |
|  | Kobayashi et al., 2017 [15] | Modified NOS | Moderate |
|  | Magill et al., 2012 [16] | Modified NOS | Moderate |
| Working with symptoms (suspected or subclinical) | Hoang Johnson et al., 2021 [42] | Modified NOS | Moderate |
|  | Ahmed at al., 2020 [13] | Modified NOS | High |
|  | Kuster et al., 2021 [59] | NOS | Moderate |
|  | Machida et al., 2020 [52] | Modified NOS | Moderate |
|  | Mitchell and Coatsworth, 2020 [61] | CASP | High |
|  | Miwa et al., 2020 [44] | Modified NOS | Low |
|  | Podewils et al., 2020 [43] | Modified NOS | Moderate |
|  | Cowman et al., 2019 [17] | Modified NOS | Moderate |
|  | Jiang et al., 2019 [40] | NOS | Moderate |
|  | Wilson et al., 2019 [18] | Modified NOS | Low |
|  | Yu et al., 2019 [45] | Modified NOS | Moderate |
|  | McGregor et al., 2018 [65] | Modified NOS | Low |
|  | Chiu et al., 2017 [19] | Modified NOS | Moderate |
|  | Mossad et al., 2017 [20] | Modified NOS | Moderate |
|  | Piper at el., 2017 [33] | Modified NOS | High |
|  | Rebmann et al., 2016 [41] | Modified NOS | Moderate |
|  | Norton et al., 2015 [23] | Modified NOS | Low |
|  | De Perio et al., 2014 [22] | Modified NOS | Moderate |
|  | Bhadelia et al., 2013 [24] | Modified NOS | Moderate |
|  | Esbenshade et al., 2013 [25] | Modified NOS | Low |
|  | Rebmann et al., 2013 [26] | Modified NOS | Low |
|  | Wang et al., 2012 [51] | Modified NOS | Low |
|  | Martinez and Ferreira, 2012 [56] | Modified NOS | Low |
|  | Jena et al., 2012 [39] | Modified NOS | Low |
|  | Askarian et al., 2011 [46] | Modified NOS | Moderate |
|  | Rousculp et al., 2010 [27] | NOS | Moderate |
|  | Iverson et al., 2010 [57] | Modified NOS | Low |
|  | May et al., 2010 [28] | Modified NOS | Low |
|  | Gudgeon et al., 2009 [29] | Modified NOS | Low |
|  | Nichol et al., 2009 [30] | NOS | Moderate |
|  | Ji et al., 2008 [49] | Modified NOS | Low |
|  | Ablah et al., 2008 [31] | Modified NOS | Moderate |
|  | Fernando et al., 2008 [47] | Modified NOS | Low |
|  | LaVela et al., 2007 [32] | Modified NOS | Moderate |
| Going to work with history of exposure to an RID | De Perio et al., 2012 [33] | Modified NOS | Low |
|  | Palmer et al., 2010 [34] | NOS | Moderate |
| Intention to attend work with an RID | Babcock et al., 2020 [35] | CASP | Moderate |
|  | Dyal et al., 2020 [14] | CASP | Moderate |
|  | Li et al., 2020 [48] | Modified NOS | Low |
|  | Tartari et al., 2020 [64] | Modified NOS | Low |
|  | Kaldjian et al., 2019 [36] | Modified NOS | Moderate |
|  | O’Neil et al., 2017 [37] | Modified NOS | Low |
|  | Szymczak et al., 2015 [38] | Modified NOS | Low |
|  | Meilicke et al., 2013 [58] | Modified NOS | Moderate |
|  | Rao et al., 2010 [50] | Modified NOS | Low |
| Adherence to guidance to stay at home from work with an RID | Brown et al., 2010 [62] | Modified NOS | Moderate |
|  | Eastwood et al., 2009 [63] | Modified NOS | Moderate |
|  | Blendon et al., 2008 [6] | Modified NOS | Moderate |

Abbreviations: *CASP* Critical Appraisal Skills Programme; *NOS* Newcastle Ottawa Scale; RID respiratory infectious disease
