## Additional file 4 for "Risk factors associated with respiratory infectious disease-related presenteeism: a rapid review"

Additional file 4: Presenteeism measurements for the included studies

| *Additional file 4* | | | | | |
| --- | --- | --- | --- | --- | --- |
|  | Reference | Indicator of presenteeism | Period of measurement | RID-presenteeism frequency | RID-presenteeism productivity |
| Working while infected (confirmed) | Jain et al., 2021 [53] | LCRC records | 8 weeks | 32.3% |  |
|  | Imai et al., 2020 [60] | Incidence and duration of sick leave taken by laboratory-confirmed employees over the observation period | 7 years | 14.1% |  |
|  | Mikos et al., 2020 [54] | Refusal to take sick leave in response to doctors’ advice. Recorded in medical records | 15 months | 26.9% |  |
|  | Juszczyk et al., 2018 [55] | Refusal to take sick leave in response to doctors’ advice. Recorded in medical records | 15 months | 32.2% |  |
|  | Kobayashi et al., 2017 [15] | Self-administered survey, interview, lab results, resident infection logs, employee absence logs | 3 months | One of six staff members worked while symptomatic |  |
|  | Magill et al., 2012 [16] | Laboratory results, interview, work schedules and medical records | 1 month | 55% |  |
| Working with symptoms (suspected or subclinical) | Hoang Johnson et al., 2021 [42] | Survey | 2017–2018 influenza season | 43.1% |  |
|  | Ahmed at al., 2020 [13] | Survey, medical records and laboratory test | 7-14 days | Mean days worked during the first 3 days of illness was 1.14 days |  |
|  | Kuster et al., 2021 [59] | Symptom diaries | Study day | 67.9% worked with symptoms of influenza infection on 8.8% of study days. |  |
|  | Machida et al., 2020 [52] | Survey | Past 1 month | 62.2% |  |
|  | Mitchell and Coatsworth, 2020 [61] | Survey | Past 2 years | 72% |  |
|  | Miwa et al., 2020 [44] | Survey | 5 months | 48.9% |  |
|  | Podewils et al., 2020 [43] | Interviews | 7 months | 82.4% (subgroup analysis: 86.4% symptomatic Hispanic and 77.3% non-Hispanic) |  |
|  | Cowman et al., 2019 [17] | Survey | Past 12 months | 54% |  |
|  | Jiang et al., 2019 [40] | Daily illness dairies | 2010–2011 to 2013–2014 influenza seasons. | 52.0% reported working on every scheduled day and 94.6% reported working at least 1 day |  |
|  | Wilson et al., 2019 [18] | Survey | 9 days | 64% |  |
|  | Yu et al., 2019 [45] | Survey | 6 months | 87% |  |
|  | McGregor et al., 2018 [65] | Survey. | Past 4 weeks |  | Mean (SD) number of days of work lost due to cold and flu-presenteeism: cold: 4.47 (8.35) and flu: 4.22 (7.38) |
|  | Chiu et al., 2017 [19] | Survey | 6.5 months | 41.4% |  |
|  | Mossad et al., 2017 [20] | Survey | Peak epidemic local influenza activity - 2 weeks in March 2016 | 93.3% with ILI and 91.7% with ILI B |  |
|  | Piper at el., 2017 [21] | 2009 MEPS linked to the 2009 medical conditions file | 2009 | nr |  |
|  | Rebmann et al., 2016 [41] | Survey | Past 3 years | 42.1% |  |
|  | Norton et al., 2015 [23] | Interview | No cut-off point. Respondents asked to recall their last experience of presenteeism | Employees - sore throat: 6.6%; cold 54.5%; cough 16.4%. Managers - sore throat: 13.6%; cold: 62.4%; cough: 12.9%; flu: 8.8% |  |
|  | De Perio et al., 2014 [22] | Survey | Past 7 months | 77% |  |
|  | Bhadelia et al., 2013 [24] | Work Health and Safety medical records | 1 year | 65% |  |
|  | Esbenshade et al., 2013 [25] | Survey | Previous influenza season | 46.1% |  |
|  | Rebmann et al., 2013 [26] | Survey | 2009 H1N1 pandemic | 14.9% |  |
|  | Wang et al., 2012 [51] | Survey | Unclear | 92.7% nurses carried on working, 57.6% self-reported colleagues as a source of infection and 59.7% infected other nurses |  |
|  | Martinez and Ferreira, 2012 [56] | Survey | 12 months | Mean [SD] number of days: 8.1 [12.2] |  |
|  | Jena et al., 2012 [39] | Survey | The prior training year | 51% of residents reported working with flu-like symptoms, and 16% reported working sick at least 3 times |  |
|  | Askarian et al., 2011 [46] | Self-administered questionnaire | Previous year | 61% |  |
|  | Rousculp et al., 2010 [27] | Monthly surveys | 6 months | 71.9% |  |
|  | Iverson et al., 2010 [57] | Survey. | Past 4 weeks |  | Mean (SD) number of workdays lost associated with cold: 4.74 (6.33) and influenza: 3.11 (4.52) |
|  | May et al., 2010 [28] | Survey | Onset of the H1N1 epidemic in April 2009 to the date of survey [November 3 and December 11, 2009] | 61% |  |
|  | Gudgeon et al., 2009 [29] | Survey | nr | 60% vs 51% vs 48% to work more than 80% of the time when ill (staff physicians vs residents vs medical students) |  |
|  | Nichol et al., 2009 [30] | Monthly surveys | The influenza season (3 months) |  | The median level of work effectiveness reported by participants for the days that they worked while ill was 70%–75% (25th percentile, 50%; 75th percentile, 80%) |
|  | Ji et al., 2008  [49] | Survey | last year | 100% |  |
|  | Ablah et al., 2008 [31] | Survey | unclear | 61% |  |
|  | Fernando et al., 2008 [47] | Survey | Previous 4 weeks |  | Mean [SD] number of days lost due to presenteeism for flu: 2.38 (8.89) and cold: 1.83 (3.48) |
|  | LaVela et al., 2007 [32] | Survey | Influenza season (5 months) | 86% |  |
| Going to work with history of exposure | De Perio et al., 2012 [33] | Survey, medical records, work documents and rotation schedule | 2 months | 77% |  |
|  | Palmer et al., 2010 [34] | Monthly surveys. | 6 months |  | Mean (SD) number of hours: 1.8 (4.3) |
| Intention to attend work with an RID | Babcock et al., 2020 [35] | nr | nr |  |  |
|  | Dyal et al., 2020 [14] | nr | nr |  |  |
|  | Li et al., 2020 [48] | Survey | No cut-off point -hypothetical question | 82.6% total. Of which, farmers: 87.6%, traders: 61.9%, trade workers: 62.5% |  |
|  | Tartari et al., 2020 [64] | Survey | Past 2 years | 58.5% total. Of which, HCWs: 56.2%, non-HCWs: 60.6% |  |
|  | Kaldjian et al., 2019 [36] | Survey | No cut off point – hypothetical question | cold: resident physicians 100%, faculty physicians 92%; flu: resident physicians 67%, faculty physicians 14% |  |
|  | O’Neil et al., 2017 [37] | Survey | No cut off point – hypothetical question | Q1. If I get a respiratory infection, I stay home until I feel better: 42%; Q2. If my coworkers get a respiratory infection they stay home until they feel better: 43.8% |  |
|  | Szymczak et al., 2015  [38] | Survey | No cut off point – hypothetical question | 55.6% |  |
|  | Meilicke et al., 2013 [58] | Two telephone surveys. The initial telephone survey was held before the influenza A H1N1 pandemic of 2008. The second survey was after the peak of the pandemic of 2009 | No cut off point – hypothetical question | 50.8% in 2008 and 40.9% in 2009 |  |
|  | Rao et al., 2010 [50] | Survey | Unclear | 24.9% |  |
| Adherence to guidance to stay at home from work with an RID | Brown et al., 2010 [62] | Survey | No cut off point – hypothetical question | Common cold: 50.7%, Seasonal influenza: 63.8%, Pandemic H1N1 2009: 96.1%, Avian influenza: 96.6% |  |
|  | Eastwood et al., 2009 [63] | Interview | No cut off point – hypothetical question | 94.1% would stay at home for 7-10 days after contact with influenza before the researchers briefly described pandemic influenza to the interviewees. 97.5% would stay at home for 7-10 days after contact with influenza after the researchers briefly described pandemic influenza |  |
|  | Blendon et al., 2008 [6] | Survey | No cut off point – hypothetical question | 94% said they would stay at home for 7–10 days if they had pandemic flu. 85% said all members of their household would stay at home for 7-10 days if a member of the same household were sick |  |

Abbreviations: *HCWs* healthcare workers; *ILI* influenza-like illness; *LCRC* London Coronavirus Response Centre; *MEPS* Medical Expenditure Panel Survey; *nr* not reported; *RID* respiratory infectious disease
