## Additional file 5 for "Risk factors associated with respiratory infectious disease-related presenteeism: a rapid review"

### Additional file 5: Reported reasons for presenteeism

| Additional file 5: Reported reasons for presenteeism | | | | |
| --- | --- | --- | --- | --- |
| **Main category** | **Sub-category** | **Study** | **Reported reasons** | **Effect measures** |
| Occupation or job | Sense of duty | Mitchell and Coatsworth, 2020 [61] | A sense of duty towards their patients (junior doctors PG year 1 and year 2) | 36% |
|  |  | Wilson et al., 2019 [18] | Sense of duty as a health care worker | 56% |
|  |  | Jena et al., 2012 [39] | Among residents who chose to work when sick, the second most frequently reported reasons an obligation to patient care | 56% |
|  |  | Chiu et al., 2017 [19] | I have a professional obligation to my coworkers | 33% (approximated from the figure) |
|  |  | De Perio et al., 2014 [22] | Feeling a professional obligation to students | 28% |
| Work and employment | Lack of cover | Miwa et al., 2020 [44] | I felt it was difficult to find replacement HCWs | 27.80% |
|  |  | Mitchell & Coatsworth, 2020 [61] | A lack of available cover (as the single biggest deterrent to taking sick leave) | 96% (32%) |
|  |  | Chiu et al., 2017 [19] | It is difficult for me to find someone to cover for me | 24% (approximated from the figure) |
|  |  | Rebmann et al., 2016 [41] | Had no one to cover for them if they were sick | 51.9% |
|  |  | May et al., 2010 [28] | No one to cover for me | 46% |
|  |  | Ji et al., 2008 [49] | Reasons for working while ill: understaffed | Qualitative assessment |
|  |  | Rebmann et al., 2013 [26] | Not having coverage by a coworker | 46% |
|  |  | Wang et al., 2012 [51] | Reasons for working while ill: not enough medical staff, too many patients, work pressure, no time to rest | Qualitative assessment |
|  | Pay loss | Dyal et al., 2020 [14] | Attendance bonuses | Qualitative risk assessment |
|  |  | Babcock et al., 2020 [35] | Potential loss of pay or vacation time | Qualitative assessment |
|  |  | Jiang et al., 2019 [40] | Could not afford to stay home | 3% |
|  |  | Wilson et al., 2019 [18] | Did not want to incur repercussions from leadership or coworkers | 20% |
|  |  | Wang et al., 2012 [51] | Reasons for working while ill: taking leave means loss of pay | Qualitative assessment |
|  | Job demand | Jiang et al., 2019 [40] | Felt miserable but had things I had to do | 11% |
|  |  | Rebmann et al., 2016 [41] | Believed they would fall behind if they did not work every day | 46.6% |
|  |  | Rebmann et al., 2013 [26] | Fear of falling behind at work | 67% |
|  | Paid Sick Leave | Hoang Johnson et al., 2021 [42] | Better sick days/PTO policy [improving absenteeism adherence] | 21% |
|  | Flexible work/leave policy | Hoang Johnson et al., 2021 [42] | Support to stay home/work from home [improving absenteeism adherence] | 12% |
| Social norms and expectations | Avoid burdening colleagues | Mitchell & Coatsworth, 2020 [61] | Concern about burdening colleagues with extra work [as the single biggest deterrent to taking sick leave] | 96% (40%) |
|  |  | Kaldjian et al., 2019 [36] | Avoid creating more work for my colleagues: resident physicians and faculty physicians | 100% and 71% |
|  |  | Babcock et al., 2020 [35] | Uncomfortable asking others to fill in for them | Qualitative assessment |
|  |  | Cowman et al., 2019 [17] | Not wanting to burden colleagues | 77% |
|  |  | Jena et al., 2012 [39] | Among residents who chose to work when sick, the most frequently reported reason was an obligation to colleagues | 57% |
|  | Peer pressure | Hoang Johnson et al., 2021 [42] | Support from coworkers [improving absenteeism adherence] | 16% |
|  |  | Hoang Johnson et al., 2021 [42] | Support from management [improving absenteeism adherence] | 14% |
|  |  | Mitchell and Coatsworth, 2020 [61] | Judgement from colleagues [as the single biggest deterrent to taking sick leave] | 44% (12%) |
|  |  | Kaldjian et al., 2019 [36] | Avoid being seen as lazy or weak: resident physicians and faculty physicians | 89% and 40% |
|  |  | Rebmann et al., 2013 [26] | Feeling pressured by a colleague[s] or supervisor to work | 36% |
|  |  | May et al., 2010 [28] | Afraid of appearing weak | 18% |
| Organisational factors | Disciplinary action or disapproval | Hoang Johnson et al., 2021 [42] | Change policy to reduce fear of disciplinary action [improving absenteeism adherence] | 13% |
|  |  | Kaldjian et al., 2019 [36] | Avoid negative repercussions: resident physicians and faculty physicians | 84% and 25% |
|  | Presenteeism culture | Babcock et al., 2020 [35] | HCP had a perception that they were encouraged to work while ill | Qualitative assessment |
|  |  | Mitchell and Coatsworth, 2020 [61] | Seeing other colleagues working when similarly unwell | 50% |
|  |  | Jiang et al., 2019 [40] | Felt miserable but felt obligated to work | 8% |
|  | Career development | Mitchell and Coatsworth, 2020 [61] | Concerns about registration requirements [as the single biggest deterrent to taking sick leave] | 48% (16%) |
| Health behaviour or perception | Not sick enough or considered a minor disease | Miwa et al., 2020 [44] | I did not feel sick enough to take sick leave; | 40.0% |
|  |  | Mitchell and Coatsworth, 2020 [61] | Not feeling sick enough to take leave | 52% |
|  |  | Wilson et al., 2019 [18] | Viewed illness as too minor to pose risk to others | 44% |
|  |  | Jiang et al., 2019 [40] | Symptoms were mild and felt well enough to work | 69% |
|  |  | Chiu et al., 2017 [19] | I could still perform my job duties; I wasn’t feeling bad enough to miss work | 47%; 38% (approximated from the figure) |
|  |  | Rebmann et al., 2016 [41] | Would work while feeling ill if their primary care provider cleared them for work; would work while feeling ill if they believed they had a mild illness | 84.2%; 74.4% |
|  |  | Rebmann et al., 2013 [26] | Belief that H1N1 pandemic influenza was a mild disease | 67.1% |
|  |  | May et al., 2010 [28] | Didn’t feel that sick | 46.0% |
|  |  | Wang et al., 2012 [51] | Reasons for working while ill: upper respiratory tract infection is considered a common disease, no need to be treated; believed they had medical knowledge and could self-treat | Qualitative assessment |
|  |  | Ji et al., 2008 [49] | Reasons for working while ill: a minor disease; can self-treat | Qualitative assessment |
|  | Lack of awareness | Chiu et al., 2017 [19] | I did not think I was contagious or could make other people sick | 35% (approximated from the figure) |
|  |  | De Perio et al., 2014 [22] | Not believing that their illness was contagious | 23.0% |
|  | Practicality | Szymczak et al., 2015 [3] | Expecting that staff will take sick leave until symptoms of upper respiratory tract infection resolve is unreasonable when resolution can take many days | Qualitative assessment |

Abbreviations: *HCP* healthcare personnel, *HCWs* healthcare workers, *PTO* paid time off
