## Additional file 6 for "Risk factors associated with respiratory infectious disease-related presenteeism: a rapid review"

*Additional file 6: Risk factors for presenteeism*

| ***Additional file 6: Risk factors for presenteeism*** | | | | |
| --- | --- | --- | --- | --- |
| **Main category** | **Sub-category** | **Study** | **Risk factors** | **Effect measures** |
| Occupation or job | Occupation | Jain et al. 2021. [53] | OR: Office work was not associated with workplace attendance after COVID-19 symptom onset | OR = 1.12, 95% CI, 0.35–4.00, P = 0.78 |
|  |  | Jain et al., 2021 [53] | OR: Retail work was not associated with workplace attendance after COVID-19 symptom onset | OR = 1.39, 95% CI, 0.41–4.74, P = 0.60 |
|  |  | Jain et al., 2021 [53] | OR: Construction work was not associated with workplace attendance after COVID-19 symptom onset | OR = 0.30, 95% CI, 0.04–2.36, P = 0.25 |
|  |  | Hoang Johnson et al., 2021 [42] | OR: Compared to other clinical roles, medical assistants were more likely to not stay home while ill in the unadjusted regression model | OR = 1.86, P = .02 |
|  |  | Imai et al., 2020 [60] | aRR: of sick leave during to lab-confirmed influenza season: Nursing, health practitioners and nonclinical professional are more likely to take sick leave than medical doctors | aRR = 1.15, 1.11, 1.13 respectively (all P < .05) |
|  |  | Imai et al., 2020 [60] | Nursing and HPs had longer sick leave than medical doctors (estimated changes (%) in sick hour rates during the periods of laboratory-confirmed infection) | 23.8% and 27.7% longer respectively (both P < .01) |
|  |  | Machida et al., 2020 [52] | OR: being a company employee (went to work within 7 days after symptom onset, compared to self-employed, part-time and government workers) | OR = 25.81, 95% CI: 2.23-298.31, P < .05 |
|  |  | Jiang et al., 2019 [40] | aRR: of working during ARI episodes was higher for physicians and lower for nurses than for other HCWs | Physicians: aRR, 1.11; 95% CI, 1.04–1.19, Nurses: aRR = 0.88; 95% CI, 0.84–0.93 |
|  |  | Bhadelia et al., 2013 [24] | Physicians and nurses were equally likely to work while symptomatic | 66.7% vs 62.5% (P = .81) |
|  |  | Ablah et al., 2009 [31] | Odds: Health care workers were more likely than other workers to report previously working while ill | 1.39 times, P <0.05 (however, the full model explained only 5% of the variance in responses) |
|  |  | Gudgeon et al., 2009 [29] | Staff physicians were the most likely to work more than 80% of the time, compared to residents and medical students (when ill with an RTI) | 60% vs 51% vs 48%, P = 0.05 |
|  |  | Gudgeon et al., 2009 [29] | Staff physicians considered the risk of transmitting that infection to others to be the lowest (compared to medical students and residents) | No figure given (P <0.001) |
|  |  | Gudgeon et al., 2009 [29] | Medical students and residents were significantly less likely than staff physicians to agree (when asked if they believed in the legitimacy of a colleague’s sick day) | 45% vs 49% vs 79%, P = 0.001 |
|  |  | Gudgeon et al., 2009 [29] | Residents were more likely than medical students or staff physicians to feel annoyed when a colleague was absent because of illness | 22% vs 11% vs 6%, P = 0.001 |
|  |  | Gudgeon et al., 2009 [29] | Surgical residents and staff were more likely than their internal medicine counterparts to come to their clinical duties more than 80% of the time when ill with an RTI | 77.5% vs 49.8%, P = 0.001 |
|  |  | Gudgeon et al., 2009 [29] | Surgeons reported a higher threshold for staying at home than internists | 8.48 vs 7.55 (on a scale of 1–10 where 5 = visibly ill), P = 0.001 |
|  |  | Gudgeon et al., 2009 [29] | Surgeons were less willing to cover a colleague’s workload rather than having them come to work when sick | 74.7% vs 88.70%, P = 0.001 |
|  |  | Gudgeon et al., 2009 [29] | Surgical residents and staff were less likely to be pleased that a sick colleague stayed at home | 75.90% vs 90%, P = 0.001 |
| Work and employment | Lack of cover | Hoang Johnson et al., 2021 [42] | OR: perceiving that their unit was understaffed (nonadherence to absenteeism among clinical and non-clinical HCWs) | OR = 1.78; P = .04 |
|  |  | Hoang Johnson et al., 2021 [42] | OR: being unable to find a replacement for work (nonadherence to absenteeism among clinical and non-clinical HCWs) | OR = 2.26; P = .03 |
|  | Paid Sick Leave |  |  |  |
|  |  | Hoang Johnson et al., 2021 [42] | OR: being paid by the hour or being unable to afford being absent (nonadherence to absenteeism among clinical and non-clinical HCWs) | OR = 2.05; P = .02 |
|  |  | Hoang Johnson et al., 2021 [42] | OR and aOR: Non-adherence to absenteeism among clinical HCWs: Did not get paid sick days | OR=2.01, 95% CI 1.26–3.36, p=.004; aOR=1.86, 95% CI 1.1–3.2, P = .02 |
|  |  | Hoang Johnson et al., 2021 [42] | When comparing HCWs who worked with ILI symptoms to those who did not: not receiving paid sick days | 11% vs 6%; P = .001 |
|  |  | Ahmed et al., 2020 [13] | AR: adjusted ratio of days worked: participants who had access to paid leave were significantly less likely to work during the first 3 days of illness | 0.81, 95% CI, 0.68–0.96, P <0.05 |
|  |  | Ahmed et al., 2020 [13] | AR: adjusted ratio of days worked at the usual workplace, participants who had access to paid leave were significantly less likely to work at usual workplace during the first 3 days of illness | 0.81, 95% CI, 0.67–0.96, P <0.05 |
|  |  | Jiang et al., 2019 [40] | aRR: Participants without paid sick leave are significantly more likely to choose "Can't afford to stay home" | aRR = 8.93; 99.9% CI, 4.20– 18.99 |
|  |  | Piper et al., 2017 [21] | APR: Employees with PSD had a higher probability of staying home for their own ILI than those without PSD | APR = 1.42, P <0.001 |
|  |  | Piper et al., 2017 [21] | APR: controlling for age, income, and education. Controlling for demographic factors, employees with PSD had a higher probability of staying home for their own influenza | APR = 1.29, P = 0.021 |
|  |  | Piper et al., 2017 [21] | PR: employed parents with PSD had a higher prevalence of staying home for a child's ILI compared to parents without PSD | PR = 1.27, P = 0.001 |
|  | Flexible work/leave policy | Ahmed et al., 2020 [13] | AR: persons with access to telework were significantly more likely to work during the first 3 days of illness | 1.25, 95% CI 1.07–1.46, P <0.01 |
|  |  | Ahmed et al., 2020 [13] | During the first 3 days of illness, the proportion who did not work at all: with access to telework vs without | 28% vs 41%, (P <0.001) |
|  |  | Ahmed et al., 2020 [13] | The mean of the total days worked: with access to telework vs without | 1.46 vs. 1.09 days, P <0.001 |
|  |  | Ahmed et al., 2020 [13] | More days off because of illness (mean): without access to telework vs with access | 1.10 vs. 0.80 days; P <0.001 |
|  |  | Machida et al., 2020 [52] | OR: unable to work from home (went to work within 7 days after symptom onset) | OR = 4.22, 95% CI: 1.02-17.43, P <0.05 |
|  |  | Rousculp et al., 2010 [27] | APP: adjusted predicted probabilities: probability that an employee attends work while their ILI symptoms are most severe, Policy: Can work from home, without policy vs with policy | 71.9% vs 57.4%, effect: -0.145, P = 0.006 |
|  |  | Rousculp et al., 2010 [27] | IRR: number of days that employees attended work while their ILI symptoms were most severe, Policy: Can work from home | IRR = 0.703, P = 0.026 |
|  |  | Eastwood et al., 2009 [63] | OR: Predicted non-compliance: Those self-employed and unable to work from home are more likely not to comply with home quarantine following exposure to pandemic influenza, compared to those not in paid employment | Univariate 7.3 (3.0–17.5), 95% CI, multivariable 4.9 (2.3–10.5), 95% CI |
|  |  | Eastwood et al., 2009 [63] | OR: Predicted non-compliance: Wage earners and those unable to work from home are more likely not to comply with home quarantine following exposure to pandemic influenza, compared to those not in paid employment | Univariate 3.4 (1.9–6.0), 95% CI, multivariable 3.3 (2.0–5.4), 95% CI |
|  | Work characteristics | Jiang et al., 2019 [40] | aRR: Participants who worked in high-risk work areas were more likely to work while symptomatic than those from other hospital areas. | aRR = 1.15; 95% CI, 1.10–1.20 |
|  | Employment status | Imai et al., 2020 [60] | Full-time was shorter than that of part-time employees (estimated changes (%) in sick hour rates during the periods of laboratory-confirmed infection) | 18.8% shorter (P < .01) |
| Social norms and expectations | Peer pressure | Rebmann et al., 2016 [41] | OR: I feel pressure from colleagues/supervisor to work while ill with symptoms of ILI | OR = 4.8 [1.5, 15.8] P = <.01 |
| Organisatio-nal factors | Presenteeism culture | Ahmed et al., 2020 [13] | AR: Persons who worked in an organisation in which employees were discouraged from coming to work if they had influenza-like symptoms were also significantly less likely to work during the first 3 days of illness | 0.86, 95% CI 0.76–0.97, P <0.05 |
|  |  | Ahmed et al., 2020 [13] | AR: Persons who worked in an organisation in which employees were discouraged from coming to work if they had influenza-like symptoms were also significantly less likely to work during the first 3 days of illness to work at their usual workplace | 0.85, 95% CI 0.74–0.96, P <0.05 |
|  |  | Rebmann et al., 2016 [41] | χ2: School nurses who indicated that their school culture encourages staff to work while ill were significantly more likely to report that they have engaged in presenteeism | χ2 = 4.6, P < .05 |
|  |  | Hoang Johnson et al., 2021 [42] | When comparing HCWs who worked with ILI symptoms to those who did not: being directed by manager to come into work | 8% vs 13%; P = .004 |
|  |  | Hoang Johnson et al., 2021 [42] | OR and aOR: Non-adherence to absenteeism among clinical HCWs: Directed by management to come to work | OR=0.44, 95% CI 0.26–0.75, p=.002; aOR= 0.37, 95% CI 0.21–0.65, P =.001 |
|  | Organisational IPC measures | LaVela et al., 2007 [32] | OR: HCWs who indicated that their facility institutes droplet precautions were significantly less likely to work while symptomatic | OR = 0.42, 95% CI, 0.19–0.94, P = 0.034 |
|  |  | LaVela et al., 2007 [32] | OR: HCWs who indicated that their facility restricts staff movement between wards and buildings were significantly less likely to work while symptomatic | OR = 0.26, 95% CI, 0.11–0.62, P = 0.002 |
|  |  | LaVela et al., 2007 [32] | OR: HCWs who indicated that their facility restricts contact between ill patients and other patients were significantly less likely to work while symptomatic | OR = 0.32, 95% CI, 0.14–0.75, P = 0.009 |
| Sociodemo-graphic factors | Gender | Jain et al., 2021 [53] | OR: Males were 66% less likely to attend the workplace with COVID-19 symptoms, compared to females | OR = 0.34, 95% CI, 0.11–1.00, P = 0.05 |
|  |  | Mossad et al., 2017 [20] | aOR: Presenteeism rates were higher in females | aOR = 2.64 (1.23–5.71) 95% CI, P = .01 |
|  |  | Piper et al., 2017 [21] | APR: Females had a higher probability of staying home for a child's illness/injury compared to males | APR = 1.46, P <0.001 |
|  |  | Piper et al., 2017 [21] | APR: Women had a higher probability of staying home for their child's ILI even after controlling for PSD access, education, income, race/ethnicity, workplace size, and family structure | APR = 1.61, P <0.001 |
|  |  | Piper et al., 2017 [21] | APR: Women had a higher prevalence than men of staying home from work for a child's influenza after controlling for PSD access, education, income, race/ethnicity, workplace size, and the presence of an unemployed adult at home | APR = 1.40, P = 0.013 |
|  |  | Brown et al., 2010 [62] | aOR: Females were more likely than males to report anticipated compliance for both common cold and seasonal influenza | common cold aOR = 1.650; CI: 1.143-2.381; seasonal influenza aOR = 1.911; CI: 1.300-2.811 |
|  |  | Eastwood et al., 2009 [63] | OR: Predicted non-compliance: Males are more likely not to comply with home quarantine following exposure to pandemic influenza | Univariate: 2.1 (1.3–3.4), 95% CI, multivariate 2.0 (1.3–3.1), 95% CI |
|  | Age | Jain et al., 2021 [53] | OR: An age of 30-49 was not significantly associated with workplace attendance after COVID-19 symptom onset, when compared with 18-29 year olds | OR = 1.36, 95% CI, 0.55–3.33, P = 0.51 |
|  |  | Jain et al., 2021 [53] | OR: An age of >50 was not significantly associated with workplace attendance after COVID-19 symptom onset, when compared with 18-29 year olds | OR = 2.00, 95% CI, 0.64-6.29, P = 0.24 |
|  |  | Mossad et al., 2017 [20] | aOR: higher in age ≤40 years | aOR = 1.95 (1.03–3.68), 95% CI, P = .04 |
|  |  | Brown et al., 2010 [62] | aOR: People age 55 and older were also more likely to report anticipated compliance for both the common cold and seasonal influenza when compared to younger respondents. | common cold aOR = 1.542; CI: 1.002-2.372; seasonal influenza aOR = 2.316; CI: 1.431-3.749 |
|  |  | Ablah et al., 2009 [31] | Odds: age 30 or younger were more likely as those age 60 and older to report previously working while ill. | 2.745 times, p<0.001 (However, the full model explained only 5% of the variance in responses) |
|  | Location | Machida et al., 2020 [52] | OR: living in a metropolitan area (for not going to work within 7 days after symptom onset) | OR = 0.06, 95% CI: 0.00-0.79, P <0.05 |
|  | Ethnicity | Podewils et al., 2020 [43] | Worked while ill (symptomatic with COVID-19): Hispanic vs non-Hispanic | 86.4% vs 77.3%, P <0.001 |
|  |  | Piper et al., 2017 [21] | APR: Hispanics had a lower probability than non-Hispanic Whites of staying home from work for their own ILI | APR = 0.83, P = 0.002 |
|  |  | Piper et al., 2017 [21] | APR: Hispanics had a lower prevalence of staying home from work when they themselves had influenza compared to non-Hispanic Whites even controlling for access to PSD, age, income, and education | APR = 0.72, P = 0.038 |
|  |  | Piper et al., 2017 [21] | APR: Hispanics had a lower prevalence of staying home for a child's ILI than non-Hispanic Whites | APR = 0.84, P = 0.035 |
|  | Social group | Rao et al., 2010 [50] | "Take leave instead of working with ILI symptoms", compared between workers, residents and students | 24.87%, 20.22% and 52.15%, χ2 = 122.27, P <.05 |
| Health behaviour or perception | Not sick enough or considered a minor disease | Hoang Johnson et al., 2021 [42] | When comparing HCWs who worked with ILI symptoms to those who did not: feeling well enough to work | 48% vs 38%; P = .001 |
|  |  | Hoang Johnson et al., 2021 [42] | OR and aOR: Non-adherence to absenteeism among clinical HCWs: Felt well enough to work | OR=1.50, 95% CI 1.13–1.99, p=.007; aOR= 1.58, 95% CI 1.15–2.17, P = .005 |
|  |  | Rebmann et al., 2016 [41] | OR: I would work while feeling ill with symptoms of ILI if I had a mild illness | OR = 3.6 [1.4, 9.7] P = .01 |
|  | Symptoms (severity) as a risk factor | Jiang et al., 2019 [40] | aRR: participants were less likely to work as symptom severity scores increased | 0.95 per 1-point increase; 95% CI, 0.94–0.95 |
|  |  | Jiang et al., 2019 [40] | aRR: participants who reported a fever were less likely to work than no fever | 0.84; 95% CI, 0.72–0.98 |
|  |  | Jiang et al., 2019 [40] | aRR: less likely to work if respiratory symptoms were accompanied by constitutional and/or gastrointestinal symptoms | 0.92; 95% CI, 0.86–0.99 |
|  |  | Jiang et al., 2019 [40] | aRR: less likely to work on the days subsequent to illness onset | 0.87; 95% CI, 0.85–0.89 |
|  | Lack of awareness | Li et al., 2020 [48] | OR: Lacking awareness of the zoonotic risk of SI | OR = 3.80, 95% CI: 1.38–11.46 |
|  |  | Li et al., 2020 [48] | OR: Not using PPE when contacting pigs | OR = 3.59, 95% CI: 1.57–8.63 |
|  |  | Eastwood et al., 2009 [63] | OR: Predicted non-compliance, knowledge of pandemic flu, those answered “Unsure/never heard of term” are more likely not to comply with home quarantine following exposure to pandemic influenza, compared to “Yes” those answered all 4 questions correct | Univariate 2.8 (1.4–6.1), 95% CI, multivariable 3.0 (1.6–5.7), 95% CI |
|  | Disease type | Juszczyk et al., 2018 [55] | The highest percentage of refusals was recorded when diagnostic codes J01-J04 were used (represented by the dark bars). The codes refer to acute sinusitis, acute pharyngitis, acute tonsillitis as well as acute laryngitis and tracheitis | 38%, 38%, 37%, 36% (approximated from figure) |
|  |  | Palmer et al., 2010 [34] | ARI-related presenteeism (employees’ own illness), ILI vs ORI, mean (SD) hours being less productive at work | 2.5 (5.0) vs 1.1 (3.3) |
|  |  | Palmer et al., 2010 [34] | ARI-related presenteeism, (household members illness), ILI vs ORI, mean (SD) hours being less productive at work | 1.3 (3.5) vs 0.4 (1.4) |
|  |  | Palmer et al., 2010 [34] | ARI-related presenteeism, (child illness), ILI vs ORI, mean (SD) hours being less productive at work | 1.4 (3.7) vs 0.5 (1.8) |
| Vaccination | Vaccination | Chiu et al., 2017 [19] | Influenza vaccinated vs not | 44.5 vs 29.2% (P = 0.03) |
|  |  | Askarian et al., 2011 [46] | No significant difference in the proportion of nurses who had continued to work with an ILI between the vaccinated and unvaccinated groups | 54% and 53%, respectively; P = 0.916 |
|  |  | Nichol et al., 2009 [30] | Adjusted difference: Mean value for vaccinated patients minus the mean value for unvaccinated patients. Vaccination is associated with statistically significant reductions in days of working while ill | -1.45 (-2.1 to -0.79) 95% CI, P <.001 |

Abbreviations: *aOR* adjusted odds ratio, *AR* adjusted ratio, *APR* adjusted prevalence ratio, *ARI* acute respiratory illness, *aRR* adjusted relative risk, *CI* confidence interval, *HPs* health practitioners, *HCWs* healthcare workers, *ILI* influenza-like illness, *IRR* incidence rate ratio, *OR* odds ratio, *ORI* other wintertime respiratory illness, *PPE* personal protective equipment, *PR* prevalence ratio, *PSD* paid sick days, *RTI* respiratory tract infection, *SD* standard deviation, *SI* swine influenza
